# Treatment recommendations based on Network Meta-Analysis: rules for risk-averse decision-makers

**DOI:** 10.1101/2024.07.01.24309758

**Authors:** A E Ades, Hugo Pedder, Annabel L Davies, H Thom, David M Phillippo, Beatrice Downing, Deborah M Caldwell, Nicky J Welton

**Affiliations:** Population Health Sciences, Bristol University Medical School, Bristol, United Kingdom

**Author notes:** **Corresponding Author:** A E Ades, PhD, Population Health Sciences, Bristol University Medical School, 37 Whately Road Bristol, BS2 8PS, United Kingdom., (44)-7879-401-276.

**Keywords:** Network meta-analysis, decision-making, loss-adjustment, expected value, treatment ranking, GRADE

## Abstract

**Background:** The treatment recommendation based on a Network Meta-analysis (NMA) is usually the single treatment with the highest Expected Value (EV) on an evaluative function. We explore approaches which recommend multiple treatments and which penalize uncertainty, making them suitable for risk-averse decision makers.

**Methods:** We introduce Loss-adjusted EV (LaEV) and compare it to GRADE and three probability-based rankings. We define the properties of a valid ranking under uncertainty and other desirable properties of ranking systems. A two-stage process is proposed: the first selects treatments superior to the reference treatment; the second identifies those that are also within a Minimal Clinically Important Difference (MCID) of the best treatment. Decision rules and ranking systems are compared on stylized examples and 10 NMAs used in NICE Guidelines.

**Results:** Only LaEV reliably delivers valid rankings under uncertainty and has all the desirable properties. In 10 NMAs comparing between 4 and 40 treatments, an EV decision maker would recommend 4-14 treatments, and LaEV 0-3 (median 2) fewer. GRADE rules give rise to anomalies, and, like the probability-based rankings, the number of treatments recommended depends on arbitrary probability cutoffs. Among treatments that are superior to the reference, GRADE privileges the more uncertain ones, and in 3/10 cases GRADE failed to recommend the treatment with the highest EV and LaEV.

**Conclusions:** A two-stage approach based on MCID ensures that EV- and LaEV-based rules recommend a clinically appropriate number of treatments. For a risk-averse decision maker, LaEV is conservative, simple to implement, and has an independent theoretical foundation.

**Highlights:** 

**What is already known?:** A risk-neutral decision-maker should make treatment decisions based on Expected Value (EV), meaning that the single treatment with the highest expected efficacy from a network meta-analysis should be recommended, regardless of uncertainty. In practice, decision makers may recommend several treatments, and take uncertainty into account on an *ad hoc* basis.

**What is new?:** We introduce Loss-adjusted EV (LaEV) as a mechanism for risk-averse decision making, and set out desirable properties of ranking systems. We define a ranking as valid under uncertainty if a higher EV is ranked above a lower one at the same uncertainty and a lower uncertainty above a higher one at the same EV. We compare LaEV to GRADE and probabilistic rankings. Of the methods examined, only LaEV provides a valid ranking under uncertainty and has all the desirable properties.

**Implications:** For a risk-averse decision maker, LaEV is a reliable, conservative, and easy-to-implement decision metric, with an independent theoretical foundation. Adoption of a risk-averse stance might focus attention on more accurate quantification of uncertainty, and encourage generation of better quality evidence.

## 1. INTRODUCTION

In decision theory a risk-neutral decision-maker bases their recommendations on the ‘Expected Value’ (EV) of a chosen evaluation function, without consideration of uncertainty in this. The evaluation function could be:

i. a measure of treatment efficacy, for example probability of an event estimated from a network meta-analysis (NMA).
ii. Net Benefit,^1^ which is monetized lifetime health gain minus lifetime costs.
iii. or any function of health improvements and adverse events, such as Multi-Criteria Decision Analysis.^2^

The choice of EV as a decision metric is based on a substantial statistical literature^3-6^ going back to the 17^th^ century.^7^ In health economic evaluations EV is regarded as optimal at a societal level^8^ as it delivers a maximally efficient allocation of resources, known as Pareto-optimality.

Faced with multiple options, an EV decision maker should therefore recommend the single treatment with the highest EV, regardless of uncertainty.^9^ In this sense, the EV decision maker is ‘risk-neutral’. In practice, however, decision makers often recommend multiple treatments, and are influenced by the degree of uncertainty in the evidence, suggesting that they are acting as risk-averse decision-makers who have a preference for more certain outcomes. In the UK, for example, multiple treatments have been recommended by NICE (National Institute of Health and Care Excellence) in both Multiple Technology Assessments,^10,11^ and more often in clinical guidelines.^12-14^ This seems to be done on an *ad hoc* basis usually when treatments have similar efficacy, reflecting a desire to keep clinical options open in case of patient differences in efficacy or side effects, factors that are seldom included in the formal decision model.

Uncertainty has also been treated in an *ad hoc* and even ambiguous manner in NICE’s official documents. The 2022 NICE manual for health technology evaluation (Section 6.3.5) requires that “the degree of certainty or uncertainty around the ICER” (Incremental Cost-Effectiveness Ratio) be taken into account.^15^ The general intention is that less should be paid for an uncertain technology (Section 6.2.34), representing a ‘risk-averse’ approach. However, if it is considered that better evidence is unlikely to be forthcoming, NICE may set a *higher* ICER threshold: this is regarded as appropriate in Highly Specialised Technology evaluations for rare diseases (Section 7.1). In this case more is paid for the more uncertain technology, representing a ‘risk-seeking’ stance. Thus, while the general decision-making position in NICE guidance is risk-neutral EV, the behaviour of NICE committees, and NICE’s own documentation, departs from EV in *ad hoc* and seemingly unprincipled ways.

Uncertainty in treatment rankings has also attracted the attention of NMA methodologists.^16-19^Besides ranking by EV, properties of alternative ‘treatment hierarchies’, or treatment rankings, have been examined formally,^20^ including: the probability of having the highest value, Pr(Best); the proportion of competitors that a treatment is superior to, also known SUCRA (Surface Under the Cumulative Ranking curve),^21^ or its equivalent the P-Score.^22^ The probability that the value of the evaluative function exceeds a certain threshold, abbreviated here as Pr(V>T), has also been studied.^23,24^

It has been proposed that these and other^25^ probability-based metrics, which, unlike EV, take uncertainty into account, could help guide NMA treatment decisions.^20,26,27^ However, by themselves ranking metrics do not define how many – or even if any – of the top-ranked treatments should be recommended. In an EV context, this can be addressed by a two-stage approach, suggested in earlier work on threshold analysis.^28^ The first stage identifies treatments that are superior to a standard reference treatment, the second selects all those that are also within a Minimal Clinically Important Difference (MCID) margin of the best treatment. The GRADE Working Group adopted a similar scheme: in Stage 1 it picks out treatments where Pr(V>T) exceeds a standard probability criterion such as 0.975.^29^ Stage 2 identifies a subset of these treatments none of which are better than any other, on the same criterion.

It has been said that “each ranking metric … answer[s] a specific treatment hierarchy question, and … every ranking metric provides a valid treatment hierarchy for the corresponding question,”^20^ a sentiment repeated in subsequent papers.^24,27,30^ However, any number of rankings and decision schemes could be proposed: we therefore need to ask: what are the properties that would make a ranking “valid” under uncertainty? And what is the “treatment hierarchy question” that decision makers *should* be trying to answer? After all, both Pr(Best) and SUCRA can have the perverse effect of privileging treatments with more uncertain effects.^18-20^

In this paper we attempt to identify and evaluate an alternative to EV which provides a rational approach to multiple treatment recommendations, and at the same time penalizes uncertainty. We will propose a metric based on Bayesian statistical decision theory,^31,32^ in which the Expected Loss arising from taking a decision under uncertainty is subtracted from the EV: we call this Loss-adjusted Expected Value (LaEV).

We begin by defining three ranking and three decision methodologies, and illustrate their properties through stylized examples. We define the validity of ranking systems under uncertainty, and suggest some desirable properties. The methodologies are then applied to ten NMAs conducted by NICE guideline developers and published in NICE guidelines and associated publications.

## 2. METHODS: DECISION RULES AND RANKING SYSTEMS

In this section we outline a range of existing decision rules and ranking systems, and propose a new metric, LaEV. We begin by defining the standard risk-neutral EV approach, and a 2-stage extension that allows for multiple recommendations. We then define the GRADE method for a ‘minimally contextualised framework’,^29^ followed by LaEV. Finally, we define three probability-based ranking systems, all familiar from previous literature, and present a 2-stage version of these so that they can be used as decision rules, and to facilitate comparisons with the other methods.

### 2.1 The NMA model and its relation to decision outcomes

We assume a standard reference treatment 1, and ‘new’ treatments 2&*k*&*K*. The NMA estimates (*K* −1) relative treatment effect parameters: {*δ*_2_&*δ*_*k*_&*δ*_*K*_} defined on the linear predictor scale. Given an estimate of the outcome on the reference treatment, *μ*, we can obtain estimates of the absolute efficacy: *μ* + *δ*_*k*_ for all *K* treatments. By convention *δ*_1_ = 0. We assume here that the joint probability distribution of these parameters is informed by a Bayesian or frequentist NMA, along with a baseline model for the target population.^37^ The link function *H* (*μ*,*δ*_*k*_) maps the linear predictor parameters onto values on the natural scale. For example, for a continuous outcome *H* (*μ*,*δ*_*k*_) = *μ* + *δ*_*k*_, and for a probability outcome *H* (*μ, δ*_*k*_) = logit^‐1^(*μ* + *δ*_*k*_).

### 2.2 Risk-neutral Expected Value

#### 2.2.1 Expected Value (Stage 1)

At Stage 1 *F*_1_(*μ*,*δ*_*k*_) − *H*(*μ*,*δ;*_*k*_) − *H*(*μ*,*δ* _1_) reflects differences between each treatment and the standard treatment 1 on the natural scale. Treatments are selected which are expected to be better than treatment 1, that is if *EV*_1_(*k*) > 0, where. 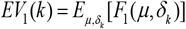.The best treatment is 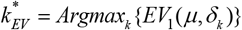. Note that throughout we assume that *F*_1_() measures the “good”outcome, so 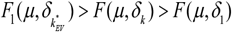. All treatments where *EV*_1_(*k*) > 0 are then considered in Stage 2.

#### 2.2.2 Expected Value (Stage 2)

The Stage 2 evaluative function compares each treatment to the best treatment and sets the difference against a threshold 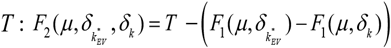. This function increases toa maximum value at *T* as treatment *k* approaches the best treatment in efficacy, and becomes negative if *k* is worse than 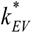 by more than *T*. *T* is therefore the maximum amount by which *k* can be inferior to 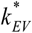 and still be recommended. The evaluative metric is the expectation over all uncertain parameters:

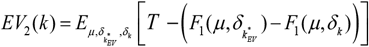

The decision rule is: adopt *k* if *EV*_1_(*k*) > 0 and *EV*_2_ (*k*) > 0.

The MCID is a natural choice for *T*, as suggested in earlier work on threshold analysis^28^ and by the GRADE Working Group.^29^

For probability outcomes, the threshold MCID might be expressed as the maximum Relative Risk by which 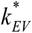 can exceed *k*. Under these circumstances, 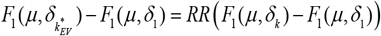 and therefore 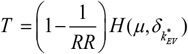 Values like 1.25 would be typical.^33^ Note that the estimated expected?value of 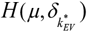 is treated as a constant for this purpose. Equivalent transformations will be required if the MCID is expressed as a hazard ratio, odds ratio, log odds ratio, or probit difference.

### 2.3 Loss-adjusted Expected Value

The decision rules based on our proposed LaEV metric are also two-staged, and parallel the rules for EV.

#### 2.3.1 LaEV Stage 1

Under uncertainty, the expected value of a (Stage 1) decision to adopt treatment *k* rather than the reference treatment on current evidence is *EV*_1_(*k*). If however we knew the parameters *μ*,*δ*_*k*_ exactly, then we would be able to select whether treatment 1 or *k* is the best treatment based on *F*_1_(*μ*,*δ* _*k*_). Over the values where *F*_1_(*μ*,*δ* _*k*_) is positive treatment *k* is best, and so there is no loss to adopting treatment *k* over treatment 1. However, over the region where *F*_1_(*μ*,*δ*_*k*_) is negative, we would obtain a higher payoff, − *F*_1_ (*μ*,*δ*_*k*_), if we adopted the current standard treatment. The Expected Loss *EL*_1_(*k*) from selecting treatment *k* rather than the reference treatment is therefore:

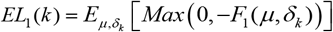

We now define the Loss-adjusted Expected Value by subtracting *EL*_1_(*k*) from *EV*_1_(*k*) :

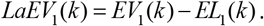

The Expected Loss of making a decision under uncertainty is equivalent to the EV of a decision made with perfect information (EVPI), an established concept in Bayesian decision theory.^31^ However, these concepts usually refer to the value of decisions between multiple treatments, whereas here interest is focussed on the value of a decision to choose a single treatment over the reference. Like the EV, the LaEV of each treatment is therefore independent of the value of the others.^34^

#### 2.3.2 LaEV Stage 2

As with EV, we introduce Stage 2 to prevent recommending treatments that are worse than the best treatment, 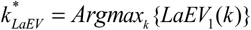, by too large a margin. The new evaluation function, 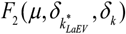 defined above for EV is applied to treatments that pass Stage 1. The Stage 2 LaEV parallels the Stage 2 EV:

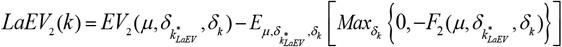

The decision rule is: adopt *k* if *LaEV*_1_ (*k*) > 0 and *LaEV*_2_ (*k*) > 0. From here, we use *k*^*^ because in all the real examples below 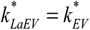, although this will not always be the case.

### 2.4 GRADE Working Group method

The GRADE two-stage process for drawing conclusions from an NMA, within what they term a ‘minimally contextualised framework’,^29^ first picks out the set of ‘Category 1’ treatments that are superior to the reference treatment, by a threshold margin *T*, with probability *P* ; in other words, all treatments *k* conforming to Pr (*F*_1_(*μ*,*δ* _*k*_) > *T*) > *P*, for example with *P* set at the standard benchmark *P* = 0.975, and *T* set to the MCID. On the second step, any Category 1 treatment *k* is promoted to Category 2 if it is superior to at least one other Category 1 treatment by the same criteria. The process continues to Category 3 or more, until we are left with a set of treatments none of which are superior to any other by the margin *T* with probability *P*. Finally, we assume that the decision rule is to recommend all treatments in the highest category. (In GRADE’s own procedures, checks for evidence inconsistency and certainty ratings may intervene before recommendations are made). The values of *T* and *P* can be changed but are assumed to stay the same within each evaluation.

### 2.5 Probability-based ranking systems

We examine three approaches: the probability of being best, Pr(best),^35^ the Surface Under the Cumulative Ranking curve (SUCRA),^21^ and the probability that the value exceeds a threshold, Pr(V>T). In the latter case the decision maker ranks treatments according to the probability that their incremental value exceeds a given threshold, *T*.^24^ The three ranking metrics are defined as follows:

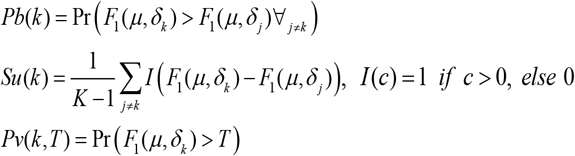

(To implement Pr(V>T) where *T* is a Relative Risk MCID, the RR is relative to reference treatment 1).

The three probability-based ranking systems are not decision rules, but the rankings can be compared to rankings generated by EV, LaEV, and GRADE. To help readers assess how they might perform as decision rules, and to aid comparison with EV-based decisions, we reported the N most highly ranked treatments in each NMA, where N is the number recommended by the EV decision rule.

## 3. Illustration of properties of ranking methods in stylized examples

In the following, we present a set of four hypothetical scenarios to illustrate, compare and contrast the properties of the alternative decision rules and ranking methods. The scenarios are explained alongside the results. WinBUGS code for each illustration is available in the Supplementary Materials.

### 3.1 Impact of uncertainty on EV, LaEV, and Pr(V>T)

Consider a one-stage two-choice decision involving relative treatment effects of a single new treatment against a standard, and evaluation functions with distribution *F*_1_(*k*) ∼ *N*(1,*σ*^2^). As we vary *σ*, there is no effect on the EV, but LaEV declines, slowly at first until *σ* is about 1, at which point it falls off in a roughly linear fashion, reaching half its value at *σ* =2.3, and turning negative at *σ* =3.6. (Fig 1a). At this point the decision maker would choose the standard treatment.

**Figure 1.**
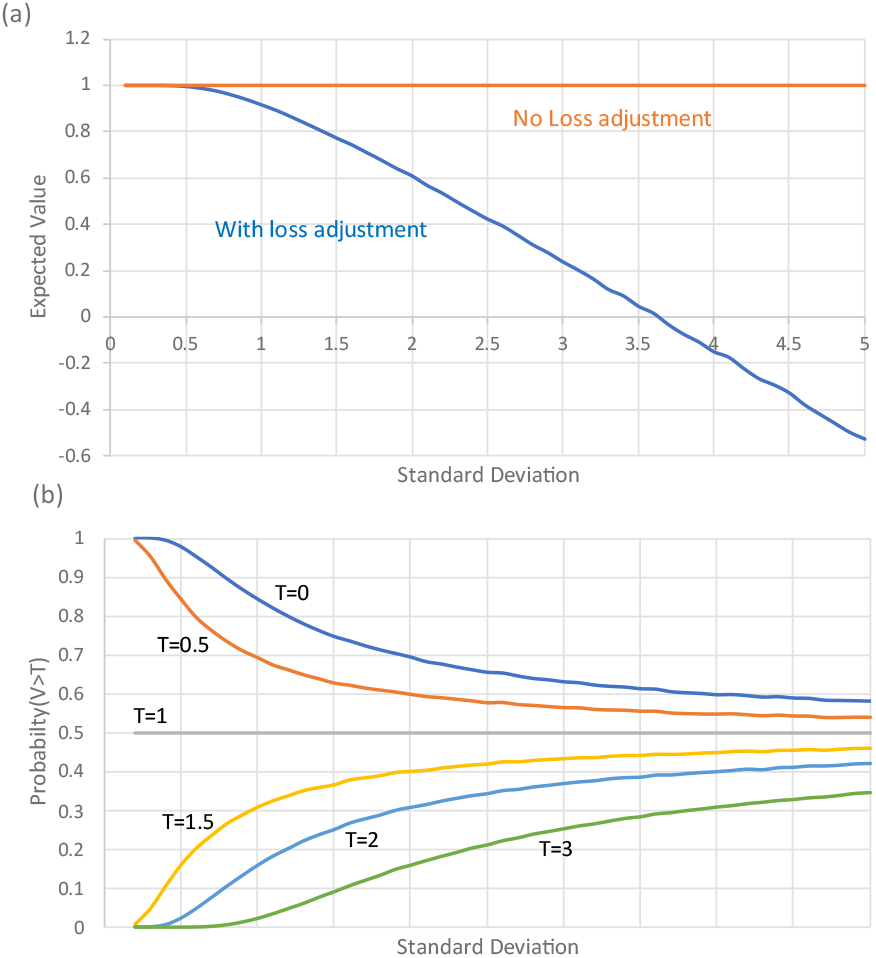
Evaluative function with mean 1.0 and SD varying from 0.1 to 5. (a) Impact of uncertainty on Expected Value with and without Loss-adjustment (b) Impact of uncertainty on Probability that the value exceeds a threshold, T.

Pr(V>T) also declines as *σ* increases, but only when T<EV. Otherwise, it rises if T>EV, or remains constant at 0.50 if T=EV (Figure 1b). Pr(V>T) therefore does not generate a ranking suitable for routine use. GRADE rules, which take the form: ‘select if Pr(V>T)>P’ are similarly limited.

#### Illustration 2: counter-intuitive properties of Pr(V>T)

Even when EV>T, Pr(V>T) can deliver counter-intuitive rankings. Figure 2 portrays the value distributions of three treatments, A,B, and C, with EVs 1.0, 2.0, 3.0. While A has the lowest EV, the uncertainty in A is negligeable, and the probability that V>0 is virtually 1. However, B and C both have an SD that is exactly one half of their EV, so the probability that V>0 is equal at 0.977. Pr(V>0) therefore ranks them (best to worst) A,B=C. In contrast, a LaEV decision maker, would rank them C,B,A with metrics (2.99, 1.99, 1.0), the same ranking as an EV-based decision, and with almost identical metrics. Metrics need to reflect the *extent* of gain or loss, not just its probability.

**Figure 2.**
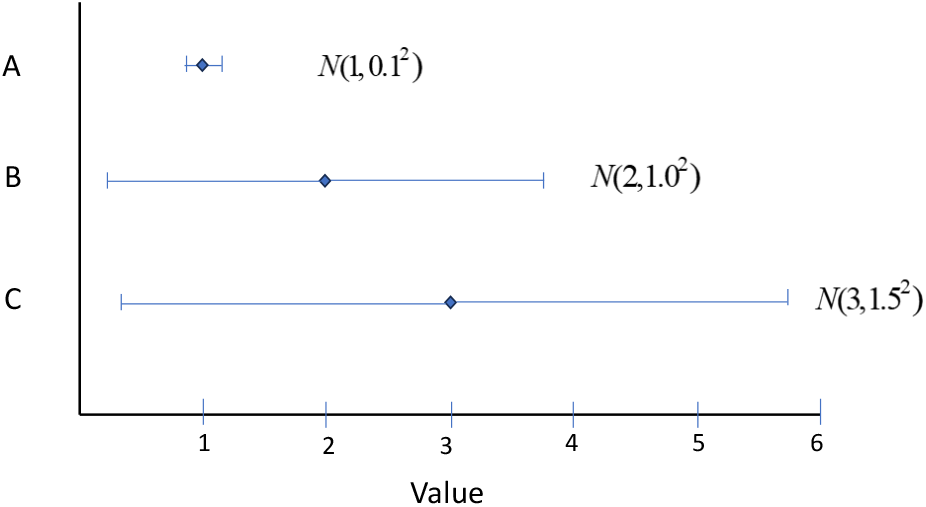
Forest plot showing Expected Value and 95% Credible intervals of three treatments, A, B, C. The probability that the value of A exceeds zero is virtually 1, while the probability that the value of B and C exceed 1 is equal at 0.977. Pr(V>0) would rank them A,B=C, with metrics (1,0.977, 0.977). An LaEV decision maker would rank them C,B,A with metrics (2.99, 1.99, 1.0), almost identical to an EV decision maker (3.0, 2.0, 1.0).

#### Illustration 3: Anomalies in GRADE decision rules

Figure 3 portrays three scenarios where GRADE rules are implemented with MCID=1 and probability threshold P=0.975. In Scenario 1 the highly uncertain treatment B is recommended along with A, while in Scenario 2, the much more certain treatment C is *not* recommended although it has the same EV as B. A treatment that reaches Stage 2 is therefore more likely to be recommended if it is uncertain.

**Figure 3.**
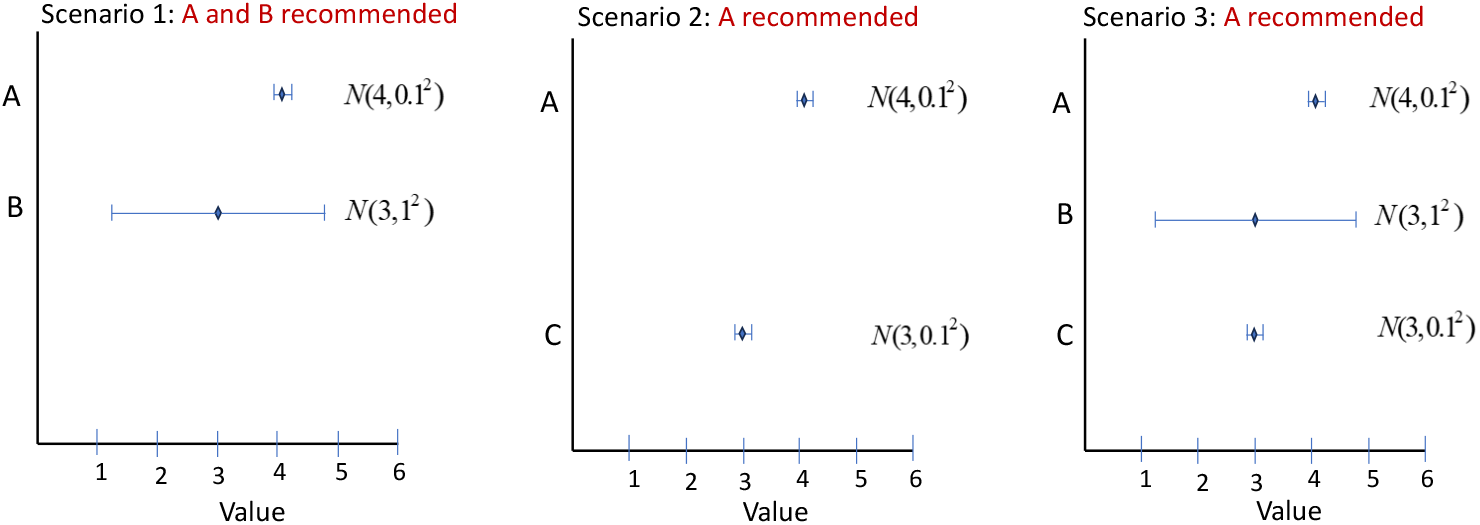
Forest plot showing Expected Value and 95% Credible intervals of three treatments, A, B, C. In Scenario 1, treatments A and B have reached GRADE Category 1 because Pr(V>1)>0.975, The MCID being 1. Because A is not superior to B by 1 with Probability 0.975, both A and B remain in Category 1 and are recommended. In Scenario 2, A is superior to C: A is promoted to Category 2 and is recommended, but C is not. In Scenario 3, A is superior to C and is promoted, while B is not.

In Scenario 3, all three treatments are compared. In contrast to Scenario 1, where both A and B are recommended, in Scenario 3 only A is recommended. The recommendation of treatment B depends on the presence or absence of treatment C, even though C would not be recommended in any of these scenarios.

#### Illustration 4: Properties of a valid ranking system in response to uncertainty

Here we consider a (one-stage) ranking of 25 treatments with evaluation functions distributed 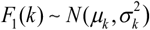 arranged in a five-by-five grid with mean *μ* = 1.1,1.2,1.3,1.4,1.5 and *σ* = 1, 2,3, 4,5. The rankings of the 25 treatments by EV, LaEV, SUCRA, Pr(Best), Pr(V>0.6), Pr(V>1.3), and Pr(V>2.3) decision rules are presented in a series of grid plots (Figure 4), in which arrows point from highest ranked treatment to the 2^nd^, then the 3^rd^, and so on. For a ranking system to be valid, the arrows must start at the lower right corner and end at the top left. Further, treatments with a higher EV must be ranked above those with a lower EV and the same SD; and those with a lower SD must be ranked above treatments with a higher SD and the same EV.

**Figure 4.**
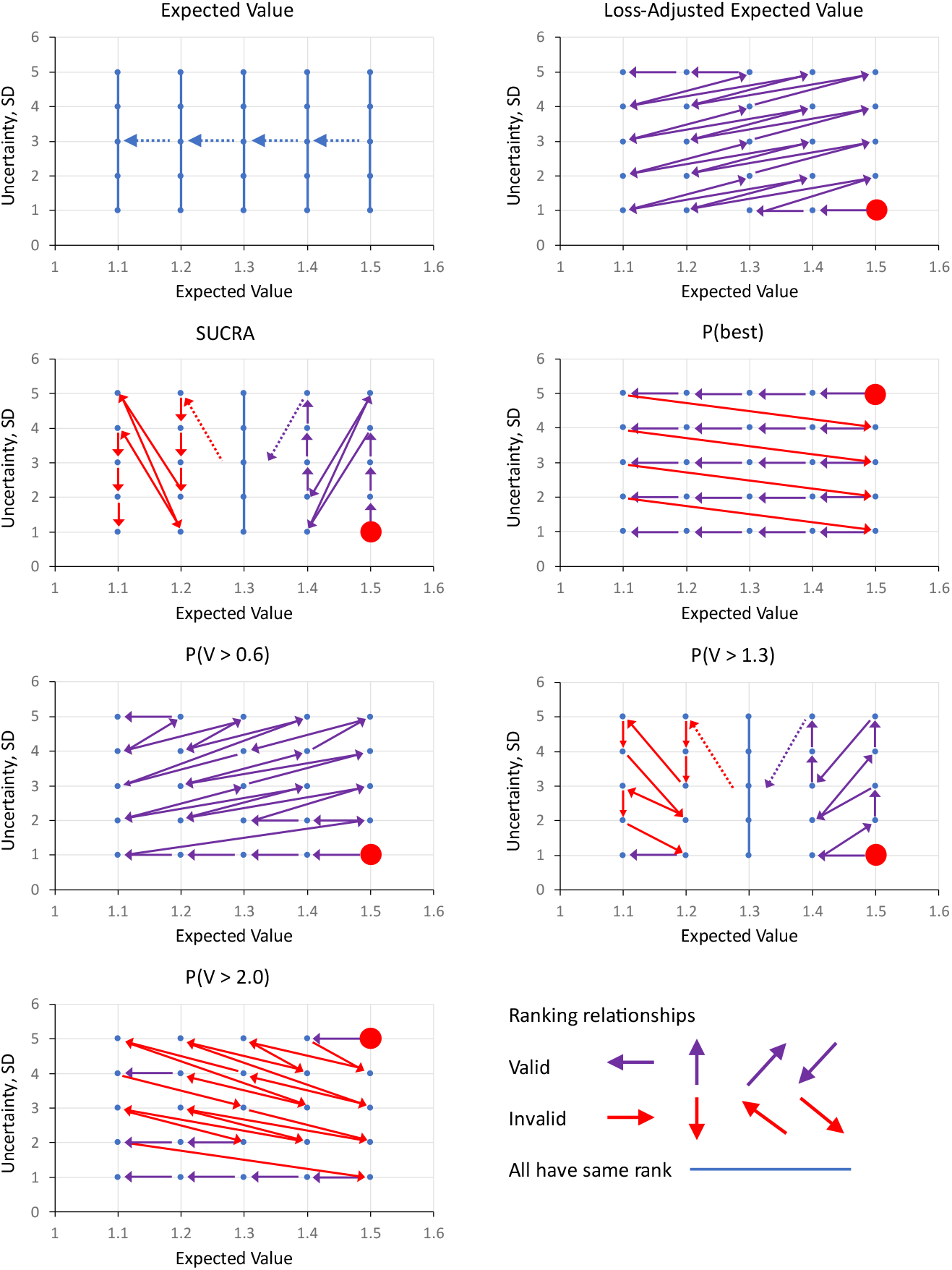
25 treatments in a 5×5 grid with EVs 1.1,1.2,1.3,1.4,1,5, and SDs 1,2,3,4,5. Rankings generated by 7 metrics: EV, LaEV, SUCRA, Pr(Best), Pr(V>0.6), Pr(B>1.3), Pr(V>2.3). Arrows start from the highest ranked treatment, marked with a red blob, and point to the 2^nd^ ranked, then the 3^rd^ ranked, and so on. Treatments linked by a blue line are of equal rank. Valid rankings (coloured purple, see Panel 8) must start at the bottom right and end at the top left. Further, they can only point Leftwards, Upwards, bottom-left to top-right, or top-right to bottom-left. Arrows pointing downwards (red) are invalid because they imply a higher ranking for a more uncertain treatment with the same EV. Likewise, arrows pointing Rightwards are invalid as they imply a higher ranking for a treatment with a lower EV at the same SD. Arrows running top-left to bottom-right imply higher ranking for treatments with both lower EV *and* higher SD. Arrows pointing bottom-right to top-left are also invalid because they skip over treatments that either have higher EV, or lower SD, or both.

Based on this simple test, EV, SUCRA, Pr(Best), Pr(V>1.3) and Pr(V>2.0) all generate invalid rankings under uncertainty. Only LaEV and Pr(V>0.6) generate exclusively valid rankings.

## 4. PREFERRED PROPERTIES AND ATTRIBUTES OF TREATMENT RANKINGS

Before turning to real examples, we summarise some preferred properties of decision rules and the treatment rankings under uncertainty, based partly on the illustrative examples. The results are set out in Table 1.

**Table 1.** Performance of alternative ranking methods regarding preferred properties. Properties marked with an asterisk are considered essential. GRADE Working Group minimally contextualized framework.; Pr(best) Probability Best; SUCRA Surface Under the Cumulative Ranking curve; Pr(V>T) probability that evaluative function exceeds threshold T; LaEV Loss-adjusted Expected Value

| Property or attribute | GRADE | Pr(best) | SUCRA | Pr(V>T) | LaEV | Comments |
| --- | --- | --- | --- | --- | --- | --- |
| Valid ranking in the face of uncertainty * | N | N | N | N | Y | Illustration 4 |
| Method should generate recommendations, not just rankings* | Y | N | N | N | Y | The probabilistic rankings by themselves do not specify how many, or even if any, treatments should be recommended |
| Methods that penalize uncertainty should only recommend as many, or fewer treatments, than EV, never more. | N | N | N | N | Y | The probabilistic rankings do not specify upper or lower limits on how many treatments would be recommended |
| Methods should not depend on arbitrary probability cutoffs | N | N | N | N | Y |  |
| Metric should be in same units as evaluation function | N | N | N | N | Y | This facilitates the use of clinically interpretable benchmarks, such as MCID |
| Must reflect extent of loss, not just its probability* | N | N | N | N | Y | Illustration 2. Also see.<sup>34</sup> |
| Metric for each treatment should be independent of the number of alternative treatments, and the value of metrics for other treatments * | ? | N | N | Y | Y | The Pr(V>T) metric underlying GRADE is independent, but its decision rule makes GRADE decisions dependent on the presence or absence of treatments that would not be recommended (Illustration 3). See<sup>34</sup> for a similar argument for independence. |
| Methods should have independent theoretical support* | N | N | N | N | Y | Only EV and LaEV have independent theoretical justifications, in the contexts of risk-neutral and -averse decision making, respectively.<sup>34,44</sup> |

## 5. RESULTS ON NICE GUIDELINES

We ran the original WinBUGS code, data, and initial values, discarding the same number of burn-in samples. Additional code generated results for decision rules and rankings (see Supplementary Materials). Results were based on at least 500,000 samples from the Bayesian posterior distribution.

### 5.1 Smoking cessation

The 2021 NICE Guidelines *Tobacco: prevention of uptake, promoting quitting and treating dependence*^14^ included an NMA of 13 classes of treatments for smoking cessation against placebo. The trial outcome was the probability of cessation. The results of both Stage 1 and Stage 2 calculations appear in Table 2. Caterpillar plots (Figure 5) show the mean (EV) and uncertainty (95%CrI) in the Stage 1 and Stage 2 evaluation functions. Also shown are the LaEV of each treatment. We have applied the EV and LaEV to all treatments at both stages for illustrative purposes: in practice only treatments with *EV*_1_(*k*) > 0 and *LaEV*_1_(*k*) > 0 would go on to Stage 2.

**Table 2.**
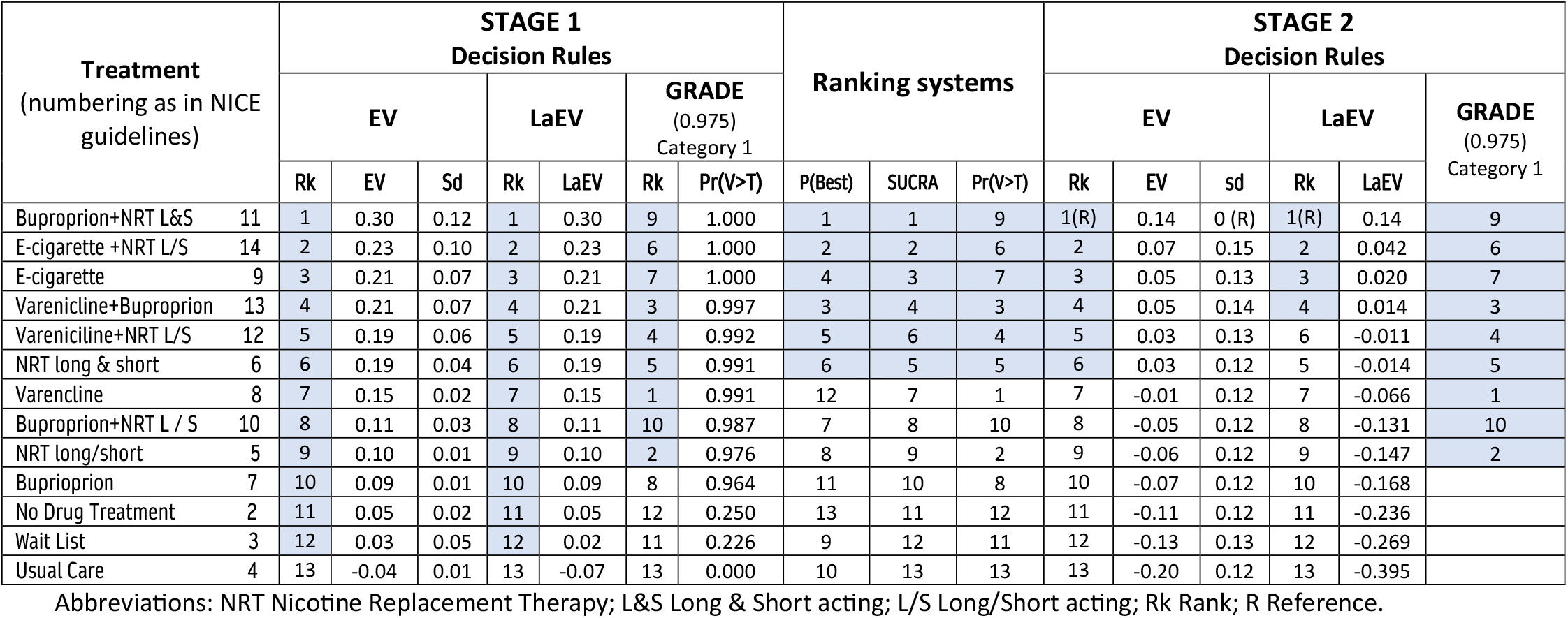
NICE Guideline Smoking Cessation.^14^ Outcome is Risk of cessation relative to Placebo. MCID based on RR=1.50. All the ranks are those generated by an EV ranking. Treatments meeting the decision criteria are shaded. For the Ranking systems in Stage 2 we have highlighted the 6 highest-rank treatments, because 6 treatments are recommended by EV.

**Figure 5.**
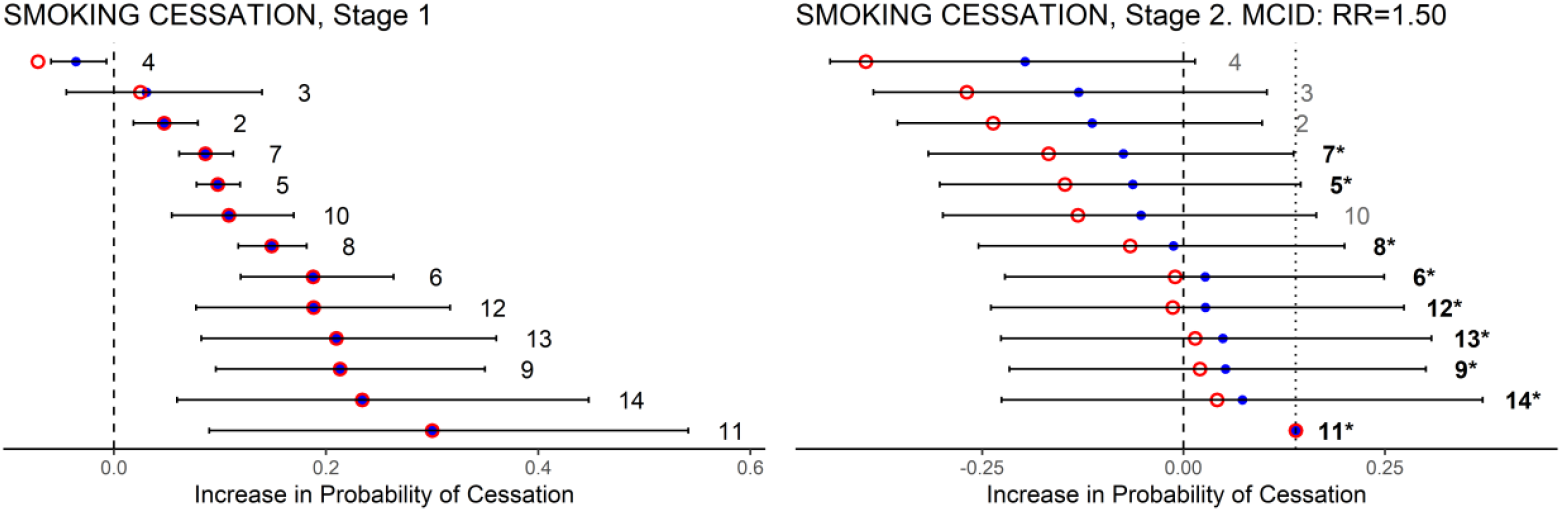
Smoking cessation. Caterpillar plots of the EV (blue dots) and its 95%CrI, and LaEV (red circles) of the Stage 1 and Stage 2 evaluation functions. Also shown: the coding of treatments in NICE guidelines; the MCID at Stage 2 (dotted line). Treatments recommended are those with EV, or LaEV, to the right of the zero (dashed) line in Stage 2. GRADE recommended treatments are those in bold and marked with asterisks.

In Stage 1, all but one of the 13 treatments could be recommended as better than placebo based on EV. Note that loss adjustment has virtually no impact on the Stage 1 valuations. This is because, although there is considerable uncertainty in the expected treatment effects, there is very little decision uncertainty: the EVs are so far from zero that the Expected Loss attaching to choosing each treatment over the reference treatment is negligeable. Accordingly, LaEV picked out the same treatments as EV (see Table 2 and Figure 3). In Stage 2, based on EV, the best treatment is joined by 5 other treatments that were not worse than the best treatment by more than the MCID (RR=1.50), while LaEV picks out 4 of these.

Application of the GRADE decision rules with the same MCID and a 0.975 cut-off resulted in 9 treatments reaching Category 1. In Stage 1, GRADE ranks treatments 9,6,7 highest because they have exceptionally low SD. Note that if a P=0.50, ‘balance of evidence’ probability had been employed, instead of 0.975, then the effect of GRADE would be identical to EV. This is what would be expected unless the distributions of the evaluative functions are highly asymmetrical. As none of the 9 Category 1 treatments were significantly better than any others by an RR of 1.50, none were promoted to Category 2, and all would therefore be recommended. However, while the ranking by GRADE at Stage 1 was quite different to the ranking by EV, at Stage 2 the 9 treatments recommended by GRADE were among the 10 most highly ranked on EV.

SUCRA delivers a ranking that is very close to EV, while the Pr(Best) ranking departs from EV quite markedly. However, if SUCRA and Pr(Best) decision makers were to recommend the same number of treatments as an EV decision maker, they would choose the same 6 treatments. A Pr(V>T) decision maker would recommend only 4 of the treatments recommended by EV.

### 5.2 Other NICE Guidelines

Detailed results, references, and commentary for a further 9 NMAs from NICE Guidelines are given in the Supplementary Materials, and all 10 are summarized in Table 3. The 10 NMAs compared between 4 and 40 treatments to the reference treatment. Some of the NMAs incorporate class models, and in some the guideline developers decided between classes of treatments. To improve network connectivity, NMA datasets sometimes include treatments that are excluded from the decision set. In these cases, we have applied rankings and decision rules only to the decision set.

**Table 3.** Summary results on 10 NMAs from NICE Guidelines. Treatment recommendations from Decision Rules (EV, LaEV, GRADE) at Stages 1 and 2, and results from ranking systems, Pr(Best, SUCRA, Pr(V>T). The numbers listed are the treatment rankings under EV. For ranking systems, the N highest ranked treatments are listed, where N is the number recommended by EV. The summary statistics for GRADE assume a 0.975 probability cutoff throughout.

| GUIDELINE | MCID | Max | Treatments recommended (number recommended) |  |  |  | Ranking systems: N Top-Ranked treatments |  |  |
| --- | --- | --- | --- | --- | --- | --- | --- | --- | --- |
|  |  |  | (N) EV | LaEV | GRADE | (P) | Pr(Best) | SUCRA | Pr(V>T) |
| **Smoking Cessation** | RR 1.5 | 13 | (6) 1-6 | (4) 1,3,2,4 | (9) 1-7,9,10 | (0.975) | 1-6 | 1-6 | 3-7,9 |
| **Moderate to Severe Acne** | CfB 25% | 26 | (14) 1-14 | (11) 1-11 | (5) 1,6-9 | (0.975) | 1-7,10,11,14,15,18,23,26 | 1-14 | 1-14 |
| **Mild to Moderate Acne** | CfB 25% | 40 | (3) 1-3 | (2) 1-2 | (1) 2 | (0.975) | 1-3 | 1-3 | 1-3 |
| **More severe Depression** | SMD 0.5 | 26 | (5) 1-5 | (3) 1,3,2 | (6) 1,3,6,7,9,10 | (0.85) | 1-5 | 1-5 | 1,3,6,9,10 |
| **Joint Replacement** | RR 1.5 | 4 | (2) 1-2 | (2) 1-2 | (1) 2 | (0.975) | 1-2 | 1-2 | 1-2 |
| **Headache** | Days 0.5 | 6 | (3) 1-3 | (3) 1-3 | (1) 3 | (0.975) | 1-3 | 1-3 | 1-3 |
| **Social Anxiety (Treatment)** | SMD 0.5 | 40 | (7) 1-7 | (5) 1-5 | (24) 1-16,19-22,24,25,27,29 | (0.975) | 1-4,6,8,28 | 1-7 | 1,2,5,7,11,19,22 |
| **Social Anxiety (Class)** | SMD 0.5 | 16 | (9) 1-9 | (6) 1-6 | (4) 1,2,5,6 | (0.975) | 1-5,7,8,10,13 | 1-9 | 1-9 |
| **Urinary incontinence** | RR 1.25 | 13 | (5) 1-5 | (2) 1-2 | (8) 1,3,6-8,10-11,13 | (0.975) | 1-4,9 | 1-5 | 6-8,11,13 |
| **Tocolytics** | Weeks 1 | 6 | (3) 1-3 | (3) 1-3 | (1) 1 | (0.975) | 1-3 | 1-3 | 1-3 |
| **SUMMARY STATISTICS** |  |  |  |  |  |  |  |  |  |
| Number recommended |  |  |  |  |  |  |  |  |  |
| Mean |  | 19 | 5.7 | 4.1 | 5.5 |  |  |  |  |
| Range (median) |  | 4-40 | 2-14 (5) | 2-11 (3) | (0-24) (2.5) |  |  |  |  |
| % of Max: Range (median) |  |  | 7-56 (38) | 5-50 (34) | (0-69) (22) |  |  |  |  |

Abbreviations: MCID=Minimal Clinically Important Difference; Max=Maximum number of treatments that could be recommended; N=number of treatments recommended by EV; P=GRADE probability cutoff; RR Relative Risk; CfB Change from Baseline; SMD Standardized Mean Difference

EV decision makers would recommend between 2 and 14 interventions (median 5), while LaEV would recommend between 3 and 11 (median 3), between zero and 3 (median 2) fewer than EV. GRADE rules with a 0.975 probability cut-off recommend between zero and 24 treatments (median 2.5), between 9 fewer and 17 more than EV. In 3/10 cases the treatment which was ranked best by EV (and LaEV) was not among the treatments recommended by GRADE. At Stage 1, GRADE privileges more certain treatments at the expense of better EV, as seen in illustration 2. However, at Stage 2, the more *un*certain treatments are recommended as they are less likely to be ‘significantly’ different from the best treatment.

The rankings produced by Pr(Best) and Pr(V>T) tend to differ from the EV and LaEV rankings, while SUCRA rankings are closer to EV. If we look at the N top-ranked treatments, where N is the number recommended by EV, SUCRA decision makers would recommend the same treatments in all 10 cases, Pr(Best) decision makers in 7, and Pr(V>T) decision makers 5.

## DISCUSSION

This paper attempts to define a rational basis for recommending more than one treatment on the basis of NMA evidence, while penalizing uncertainty. This represents a risk-averse decision-making position, in contrast to the standard EV approach. Using stylized illustrations and real examples from NICE Guidelines, we have compared EV, LaEV, and GRADE decision rules. The performance of rankings systems based on Pr(Best), SUCRA, and Pr(V>T) has also been documented, in view of the growing literature proposing that probabilistic rankings can help inform recommendations.^20,23-27,36^

We defined a ranking as valid under uncertainty if a treatment with a higher EV must always be ranked above a treatment with a lower EV and the same uncertainty, and a treatment with less uncertainty must always be ranked above a treatment with more uncertainty at the same EV. Of the methods examined only LaEV provides a valid ranking on this definition. Pr(V>T) is valid only if EV>T *for all treatments*, a property which blocks its use in routine applications, and which is inherited by the GRADE Working Group rules for a minimally contextualised framework. Although SUCRA usually generates a ranking close to EV in real examples, except when treatments differ substantially in uncertainty, it cannot be relied on to produce valid rankings under uncertainty, and it possesses none of the preferred properties. The probabilistic ranking systems and GRADE all take uncertainty into account, sometimes in irrational ways, but they do not always penalize it. They may register the probability of loss, but not its extent. Their fundamental drawback is that they do not distinguish between uncertainty in treatment effects from uncertainty in decision.

Because the EV and LaEV decision metrics are on the same scale as the evaluative function, they can access the MCID. This provides a natural basis for deciding how many treatments besides the best treatment should be recommended. MCID has been used in this way in NMA threshold analyses^28^ and has had a similar role in Bayesian sensitivity analyses more generally.^37^ Because Expected Loss is always positive (for treatments better than the reference), LaEV decision rules cannot recommend more treatments than EV, and any approach that penalizes uncertainty should have this property. The number of treatments recommended by GRADE sometimes exceeded EV, and is effectively arbitrary, subject only to the choice of probability cutoff. SUCRA delivers rankings close to the EV ranking in real examples, Pr(Best) and Pr(V>T) less so, but arbitrary cutoffs would again be required to control the number of treatments recommended by all three probabilistic ranking methods.

Adoption of any risk-averse decision rule would put a new spotlight on uncertainty and its sources. Much of the uncertainty in model parameters originates in sampling error in their estimation, but variation arising from random effects models also contributes, representing, perhaps, the uncertain relevance^38^ of evidence from trials with widely dispersed treatment effects. These sources of uncertainty are ‘within’ the decision model and can therefore engage risk-averse methods for decision making. On the other hand, the use of GRADE certainty ratings^39^ and Risk of Bias tools^40^ identifies further sources of uncertainty which tend to be treated as external or contextual factors that are ‘taken into account’ alongside the results of formal modelling. Model structure and choice of data sources represent further sources of uncertainty outside the decision model, often addressed by sensitivity analyses. Adopting decision rules that penalize uncertainty would encourage investigators to bring all such sources of uncertainty *into* the decision model, and would place a premium on statistical methods that reduce between-study heterogeneity, including: informative priors on variance parameters;^41^ bias modelling;^42^ and methods that increase precision such as multi-level network meta-regression.^43^ Bias models are already in common use in NICE guidelines.

The circumstances under which a risk-averse posture is appropriate remain a matter of debate, and beyond the scope of this paper. Briefly, an EV (risk-neutral) position is considered appropriate for a decision maker making large numbers of decisions under uncertainty,^44^ for example a national reimbursement agency. Put simply, the risks ‘average out’. However, for individual patients making a one-time decision, a risk averse stance – penalizing uncertainty – would be justified. Risk aversion is also appropriate for institutional decision makers if costs or benefits are born by individuals and cannot be transferred,^44^ or where payers have limited budgets.^34,44^ Although clinical guidelines may apply to large numbers of patients, guideline development groups typically take one-time decisions. There is empirical evidence that both patients^45-47^ and clinicians^48^ are risk averse when facing health care decisions.

A limitation of this paper is that we have not discussed other approaches to risk aversion in the literature, including: mean-variance trade-offs, methods setting a maximum probability of a poor outcome, and methods where risk aversion is a parameter input. These alternatives have seen limited uptake^34^ and none have been considered in the NMA literature. In most cases, fair comparisons would be difficult to contrive, as additional parameters are required whose values are to some extent arbitrary. A possibly more serious short-coming is our focus on risk aversion, excluding the potential role of a risk-seeking stance. Prospect Theory asserts that risk posture depends on baseline risk,^49^ and there is evidence that, in health care decisions, individuals are risk-seeking at low levels of baseline health.^46,50-53^ In the context of Net Benefit analysis this has been addressed by Generalized Risk-Adjusted Cost-Effectiveness (GRACE), in which willingness-to-pay varies with baseline risk.^54,55^ It may be, therefore, that LaEV as elaborated here is not suited to life-threatening conditions, or where the baseline life expectancy or quality-adjusted life expectancy is low. Whether our proposals can be extended to allow risk posture to depend on baseline health status, and more generally to evaluations based on Net Benefit, are topics of on-going research. LaEV appears to constitute a relatively conservative methodology for risk-averse decision makers. In the 10 examples, it recommended only 0-3 fewer (median 2) treatments than EV. It requires an SD of 2.3 units to halve a single unit of EV, and an SD of 3.6 units to entirely neutralise it (Illustration 1). We can therefore anticipate that if LaEV was to replace EV-based decision making, the impact would be no more than moderate. A more substantial impact would be expected where highly uncertain evidence is used, for example evaluations based on non-randomised evidence, or ‘unanchored’ comparisons.^56^ This underscores the importance of properly representing uncertainty within the decision model: if this was implemented, routine use of risk-averse decision-making methods might incentivize the production of better quality data,^57^ reversing the trend towards accepting evidence from non-randomised and one-arm studies.

Methods used by guideline developers need to be acceptable to key stakeholders, including professional colleges, manufacturers, health care workers, and patients. Stakeholders require a degree of certainty regarding which methods for health technology assessment are acceptable, and how they are to be applied. To achieve this, methods have to meet criteria for transparency and consistency across conditions.^58^ This weighs against methods where parameters can be set in arbitrary ways, therefore against GRADE and against decision rules based on SUCRA or Pr(V>T) rankings, if they were to be proposed. Also problematic are ranking methods that combine efficacy with other outcomes such as adverse effects, costs, or GRADE certainty ratings, using arbitrary, condition-dependent weightings,^23^ even if they were able to reliably produce valid rankings under uncertainty. More fundamentally, GRADE and the probabilistic ranking systems, and indeed other novel ranking approaches,^25,36^ stand outside the standard theory and practice of health evaluation. Indeed, no theoretical basis has been proposed in which any of these methods would represent an optimal basis for decision making.

In 2001, the Institute of Medicine identified ‘patient centred medicine’ as an objective for improved health in the 21^st^ century,^59^ and this was widely endorsed by research funders and organisations delivering health care. Patient-centric decision making was seen as an essential component. Given that individuals are generally risk averse when facing health care decisions, a risk-averse methodology by guideline developers would be a step towards patient-centred medicine. For this purpose, the two-stage LaEV method can be recommended as reliable, conservative, theoretically well-motivated, and simple to implement.

## AUTHOR CONTRIBUTIONS

**A E Ades**: Conceptualisation; analysis; software; writing - original draft; writing – review and editing. **Hugo Pedder**: conceptualisation; writing – review and editing. **Annabel L. Davies**: conceptualisation; writing – review and editing. **H Thom**: conceptualisation; writing – review and editing. **David M Phillippo**: conceptualisation; writing – review and editing. **Beatrice Downing**: writing – review and editing. **Deborah M Caldwell**: conceptualisation; writing – review and editing. **Nicky J Welton**: conceptualisation; writing – review and editing

## DATA AVAILABILITY

No new data were created or analysed in this study. The original NMA data and WinBUGs code are available from the cited guidelines.

