## Supplemental File for "Treatment recommendations based on Network Meta-Analysis: rules for risk-averse decision-makers"

A. **Detailed results on NMAs from NICE guidelines**
   - **A1: Moderate to Severe Acne**
   - **A2: Mild to Moderate Acne**
   - **A3: More Severe Depression**
   - **A4: Tranexamic Acid in Joint Replacement**
   - **A5: Headache prophylaxis**
   - **A6: Social Anxiety (Treatment)**
   - **A7: Social Anxiety (Class)**
   - **A8: Urinary Incontinence**
   - **A9: Tocolytic Therapy**

B. **WinBUGS code for illustrative examples**

C. **WinBUGS code for NICE Guidelines**

D. **References**

#### Note on the Supplementary Tables and Figures.

The tables show the results of applying the decision rules and ranking systems in a series of NMAs from Nice guidelines. All the rankings refer to the ranks generated by the EV. For the EV, LaEV, and GRADE decision rules, the shaded rankings indicate which treatments pass the Stage 1 criteria and are recommended at Stag3 2. . Note that, at Stage 2, the treatments that are top-ranked by EV and LaEV are the reference treatments, and the value of the Stage 2 evaluative function is the MCID with no uncertainty.

For the probabilistic rankings, we have shaded the N top-ranked treatments, where N is the number of treatments recommended by EV

The figures are caterpillar plots of the EV (blue dots) and its 95%CrI, and LaEV(red circles) of the Stage 1 and Stage 2 evaluation functions, ordered by EV. Also shown: the coding of treatments in NICE guidelines; the MCID at Stage 2 (dotted line). Treatments recommended are those with EV, or LaEV, to the right of the zero (dashed) line in Stage 2. GRADE recommended treatments are those in bold and marked with asterisks.

LaEV SUPP
June 18 2024

#### APPENDIX A: DETAILED RESULTS ON NMAS FROM NICE GUIDEINES

##### A1. Moderate-to-Severe Acne[1]

**Table S1**

| Treatment (numbering as in NICE guidelines) | | STAGE 1 Decision Rules | | | | | | | Ranking systems | | | STAGE 2 Decision Rules | | | | | |
|---|---|---|---|---|---|---|---|---|---|---|---|---|---|---|---|---|---|
| | | EV | | | LaEV | | GRADE (0.975) Category 1 | | | | | EV | | | LaEV | | GRADE (0.975) Category 1 |
| | | Rk | EV | Sd | Rk | LaEV | Rk | Pr(V>T) | P(Best) | SUCRA | Pr(V>T) | Rk | EV | sd | Rk | LaEV | |
| Retinoid tcd ≥ 120mg/kg (sc)[o] | 9 | 1 | 58.0 | 10.7 | 1 | 58.0 | 1 | 0.999 | 2 | 1 | 1 | 1(R) | 25.0 | 0 (R) | 1(R) | 25.0 | 1 |
| *PhTh therapy | 17 | 2 | 57.6 | 17.4 | 2 | 57.6 | 8 | 0.994 | 1 | 2 | 8 | 2 | 24.6 | 20.0 | 2 | 23.5 | 8 |
| *Nicotinamide[t] | 7 | 3 | 49.9 | 13.8 | 3 | 49.9 | 7 | 0.993 | 3 | 3 | 7 | 3 | 16.9 | 16.9 | 3 | 15.5 | 7 |
| *PhTh+photodynamic therapy | 16 | 4 | 47.9 | 15.4 | 4 | 47.9 | 9 | 0.984 | 4 | 5 | 9 | 4 | 14.9 | 18.3 | 4 | 12.7 | 9 |
| Retinoid tcd<120mg/kg (sc)[o] | 8 | 5 | 47.6 | 14.1 | 5 | 47.6 | 6 | 0.981 | 5 | 4 | 6 | 5 | 14.6 | 17.2 | 5 | 12.7 | 6 |
| *Tetracycline [oral]+PhDy therapy | 27 | 6 | 44.8 | 9.5 | 6 | 44.8 | 2 | 0.968 | 6 | 7 | 2 | 6 | 11.8 | 13.2 | 6 | 10.5 | |
| Lincosamide[t]+Retinoid[t] | 22 | 7 | 44.5 | 7.8 | 7 | 44.5 | 3 | 0.965 | 11 | 6 | 3 | 7 | 11.5 | 12.6 | 7 | 10.3 | |
| BP[t]+Retinoid[t]+Tetracycline[o] | 26 | 8 | 43.5 | 7.2 | 8 | 43.5 | 12 | 0.958 | 7 | 8 | 12 | 8 | 10.5 | 8.2 | 8 | 10.1 | |
| Photodynamic therapy | 15 | 9 | 40.5 | 7.1 | 9 | 40.5 | 5 | 0.945 | 18 | 9 | 5 | 9 | 7.5 | 12.1 | 9 | 5.5 | |
| *No treatment | 2 | 10 | 39.4 | 18.7 | 10 | 39.2 | 4 | 0.931 | 14 | 10 | 4 | 10 | 6.4 | 21.2 | 10 | 0.7 | |
| Azelaic acid[t]+Tetracycline[o] | 24 | 11 | 38.4 | 15.8 | 11 | 38.4 | 13 | 0.919 | 10 | 11 | 13 | 11 | 5.4 | 18.7 | 11 | 0.4 | |
| Retinoid[t]+Tetracycline[o] | 25 | 12 | 35.2 | 5.9 | 12 | 35.2 | 11 | 0.802 | 15 | 12 | 11 | 12 | 2.2 | 11.1 | 12 | -1.1 | |
| Lincosamide[t] | 4 | 13 | 34.1 | 6.6 | 13 | 34.1 | 14 | 0.791 | 26 | 14 | 14 | 13 | 1.1 | 11.8 | 13 | -3.0 | |
| BP[t]+Retinoid[t] | 21 | 14 | 34.0 | 11.1 | 14 | 34.0 | 10 | 0.782 | 23 | 13 | 10 | 14 | 0.9 | 15.5 | 14 | -4.7 | |
| PhCh therapy[red] | 12 | 15 | 29.7 | 11.5 | 15 | 29.7 | 16 | 0.675 | 8 | 15 | 16 | 15 | -3.3 | 15.2 | 15 | -11.1 | |
| Benzoyl peroxide[t] | 3 | 16 | 28.9 | 8.6 | 16 | 28.9 | 15 | 0.662 | 9 | 16 | 15 | 16 | -4.1 | 13.8 | 16 | -11.9 | |
| PhCh+PhTh therapy | 14 | 17 | 28.3 | 15.7 | 17 | 28.0 | 17 | 0.587 | 16 | 17 | 17 | 17 | -4.7 | 18.6 | 17 | -14.7 | |
| *Co-cyprindiol[o] | 11 | 18 | 25.0 | 15.5 | 18 | 24.7 | 18 | 0.501 | 21 | 18 | 18 | 18 | -8.0 | 18.9 | 19 | -18.6 | |
| Tetracycline[o] | 10 | 19 | 24.2 | 4.1 | 19 | 24.2 | 19 | 0.423 | 12 | 19 | 19 | 19 | -8.8 | 10.0 | 18 | -20.1 | |
| BP[t]+Lincosamide[t]+Retinoid[t] | 23 | 20 | 23.1 | 7.5 | 20 | 23.1 | 23 | 0.406 | 20 | 20 | 23 | 20 | -9.9 | 13.1 | 20 | -21.6 | |
| BP[t]+Lincosamide[t] | 19 | 21 | 22.7 | 8.4 | 21 | 22.7 | 20 | 0.394 | 17 | 23 | 20 | 21 | -10.3 | 13.6 | 21 | -22.3 | |
| BP[t]+Macrolide[t] | 20 | 22 | 22.1 | 4.8 | 22 | 22.1 | 21 | 0.391 | 13 | 21 | 21 | 22 | -10.9 | 11.8 | 22 | -22.9 | |
| *BP[t]+Anti-fungal[t] | 18 | 23 | 22.1 | 12.2 | 23 | 21.9 | 22 | 0.272 | 24 | 22 | 22 | 23 | -10.9 | 16.3 | 23 | -24.2 | |
| Retinoid[t] | 5 | 24 | 13.1 | 2.5 | 24 | 13.1 | 26 | 0.258 | 25 | 26 | 26 | 24 | -19.9 | 11.0 | 24 | -39.9 | |
| Macrolide[t] | 6 | 25 | 10.9 | 7.3 | 25 | 10.7 | 25 | 0.028 | 19 | 24 | 25 | 25 | -22.1 | 13.1 | 25 | -44.5 | |
| *PhCh therapy[blue and red] | 13 | 26 | 8.8 | 24.8 | 26 | 2.8 | 24 | 0.000 | 22 | 25 | 24 | 26 | -24.2 | 26.7 | 26 | -50.7 | |

Abbreviations: BP Benzoyl peroxide; tcd total cumulative dose; [t] topical; [o] oral; [ph] physical; (sc) single course; ToAc Topical Acid; PhCh Photochemical therapy; PhTh Photothermal therapy; PhDy Photodynamic therapy; Comb. Combined; Rk rank (values are ranks on EV)

***Moderate-to-Severe Acne***

***Type of model**:* Random study effects, fixed class effects model with 52 treatments in 26 classes relative to placebo.

***Outcome*:** % Change from Baseline.

***MCID***: 25%, ***GRADE probability cutoff**:* 0.975

***Decision rules Stage 1*** **:**
*EV:* all 26 treatment classes have positive EV
*LaEV:* all 26 treatment classes have positive LaEV
*GRADE:* 5 treatments qualify and are promoted to Category 1.

***Ranking Systems***: Pr(V>T) places treatments ranked 8,7,9,6 on EV among the top 5: these have exceptionally low SD. But both Pr(V>T) and SUCRA pick out the same top 14 treatments as EV. The Pr(Best) ranking is erratic, privileging more uncertain treatments

***Decision rules Stage 2*:**
*EV*: 13 treatments besides the best treatment are recommended, because they are not worse than the best treatment by more than the MCID.
*LaEV:* 10 treatments, besides the best treatment would be recommended.
*GRADE:* none of the 5 Category 1 treatments are significantly better than any other by the MCID. Therefore, none are promoted to Category 2, and all are recommended.

***Comments:*** During consideration of the NMA results by NICE Guidelines developers, a number of treatments were ruled out on the basis of potential bias reflected in their small sample size. These are marked with an asterisk in Table S1. Note that none of those with small sample size would have been ruled out by LaEV.

**Figure S1.** Moderate to Severe Acne.

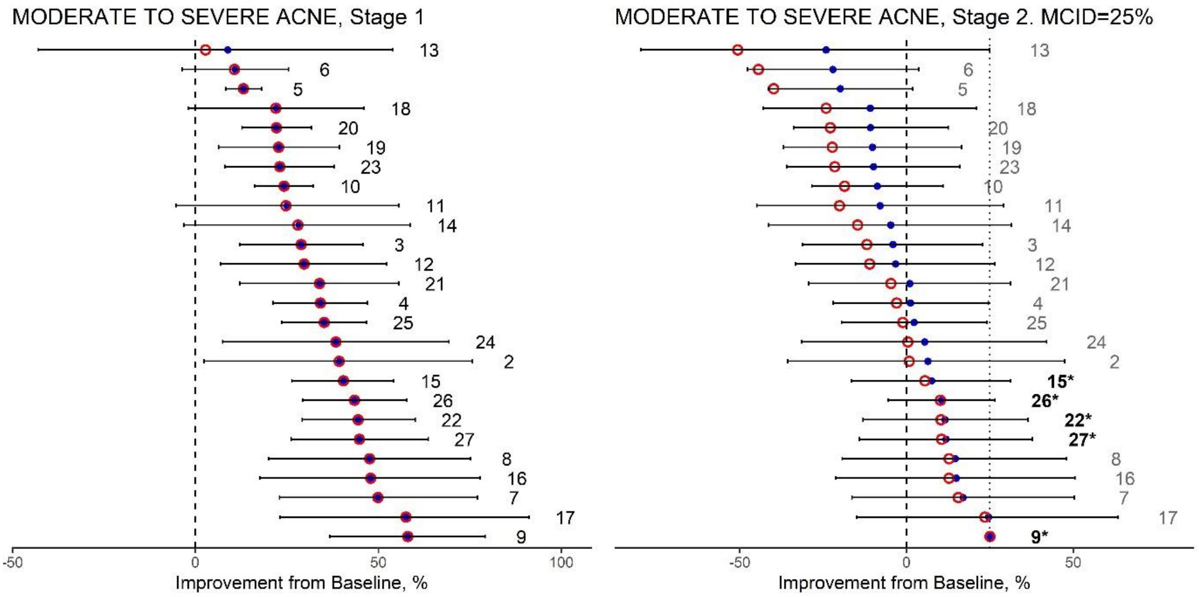

#### A2. Mild to Moderate Acne

#### Table S2

| Treatment (numbering as in NICE guidelines) | | STAGE 1 Decision Rules | | | | | | | Ranking systems | | | STAGE 2 Decision Rules | | | | | |
|---|---|---|---|---|---|---|---|---|---|---|---|---|---|---|---|---|---|
| | | EV | | | LaEV | | GRADE (0.975) Category 1 | | | | | EV | | | LaEV | | GRADE (0.975) Category 1 |
| | | Rk | EV | Sd | Rk | LaEV | Rk | Pr(V>T) | P(Best) | SUCRA | Pr(V>T) | Rk | EV | sd | Rk | LaEV | |
| *PhCh [red] | 23 | 1 | 80.8 | 40.7 | 1 | 80.4 | 2 | 0.984 | 1 | 2 | 2 | 1(R) | 25.0 | 0 (R) | 1(R) | 25.0 | 2 |
| *ACNICARE[ph] | 15 | 2 | 79.2 | 26.2 | 2 | 79.2 | 3 | 0.937 | 2 | 3 | 3 | 2 | 23.4 | 48.1 | 2 | 13.8 | |
| *PhTh+photodynamic | 26 | 3 | 64.7 | 25.9 | 3 | 64.7 | 4 | 0.933 | 3 | 1 | 4 | 3 | 9.0 | 40.1 | 3 | -2.9 | |
| *Smoothbeam+PhCh [blue] | 27 | 4 | 51.8 | 17.7 | 4 | 51.8 | 1 | 0.921 | 4 | 4 | 1 | 4 | -4.0 | 43.9 | 4 | -23.6 | |
| Chemical peel[ph] | 13 | 5 | 36.9 | 14.0 | 5 | 36.9 | 5 | 0.803 | 5 | 5 | 5 | 5 | -18.9 | 42.7 | 5 | -47.0 | |
| PhCh[b&r] | 21 | 6 | 32.6 | 9.3 | 6 | 32.6 | 6 | 0.788 | 8 | 6 | 6 | 6 | -23.2 | 41.4 | 6 | -53.9 | |
| *Photodynamic | 25 | 7 | 30.9 | 21.8 | 7 | 30.1 | 9 | 0.664 | 11 | 9 | 9 | 7 | -24.9 | 37.2 | 8 | -55.0 | |
| *No treatment | 2 | 8 | 29.5 | 33.1 | 9 | 29.5 | 10 | 0.660 | 10 | 10 | 10 | 8 | -26.3 | 27.5 | 7 | -55.4 | |
| *BP[t]+Lincosamide[t]+ToAc[t] | 40 | 9 | 29.5 | 10.6 | 10 | 29.3 | 7 | 0.613 | 9 | 12 | 7 | 9 | -26.3 | 41.8 | 9 | -59.3 | |
| *Retinoid[t]+$H_2O_2$[t] | 34 | 10 | 29.3 | 10.5 | 11 | 28.4 | 11 | 0.593 | 13 | 11 | 11 | 10 | -26.5 | 41.8 | 10 | -59.7 | |
| *Superoxidised solution[t] | 10 | 11 | 28.5 | 14.0 | 12 | 27.4 | 12 | 0.591 | 12 | 14 | 12 | 11 | -27.3 | 42.6 | 11 | -61.3 | |
| *Lincosamide[t]+Azelaic acid[t] | 31 | 12 | 27.5 | 10.2 | 13 | 26.4 | 8 | 0.572 | 6 | 7 | 8 | 12 | -28.3 | 41.6 | 12 | -62.8 | |
| *BP[t]+PhCh+PhTh | 41 | 13 | 26.5 | 11.8 | 8 | 26.0 | 13 | 0.547 | 40 | 13 | 13 | 13 | -29.3 | 42.1 | 13 | -64.7 | |
| PhCh [blue] | 22 | 14 | 25.9 | 8.5 | 14 | 25.9 | 14 | 0.534 | 25 | 15 | 14 | 14 | -29.9 | 41.1 | 14 | -65.5 | |
| BP[t]+Retinoid[t] | 30 | 15 | 23.3 | 5.2 | 15 | 23.3 | 16 | 0.416 | 27 | 16 | 16 | 15 | -32.5 | 40.8 | 15 | -69.9 | |
| *Azelaic acid[t]+Macrolide[t] | 37 | 16 | 23.1 | 9.5 | 16 | 23.1 | 15 | 0.370 | 19 | 8 | 15 | 16 | -32.7 | 41.4 | 16 | -70.4 | |
| Lincosamide[t]+Retinoid[t] | 32 | 17 | 21.4 | 7.2 | 17 | 21.4 | 18 | 0.325 | 18 | 17 | 18 | 17 | -34.4 | 41.1 | 17 | -73.5 | |
| *Macrolide[t]+Anti-fungal[t] | 3 | 18 | 20.0 | 11.5 | 18 | 19.8 | 22 | 0.316 | 7 | 18 | 22 | 18 | -35.8 | 42.0 | 18 | -76.3 | |
| *Retinoid[t]+ToAc[t]+PhCh[b&r] | 39 | 19 | 17.4 | 13.3 | 20 | 17.1 | 17 | 0.301 | 21 | 20 | 17 | 19 | -38.4 | 42.5 | 19 | -81.0 | |
| BP[t]+Macrolide[t] | 29 | 20 | 17.3 | 9.8 | 19 | 16.8 | 19 | 0.278 | 16 | 19 | 19 | 20 | -38.5 | 41.6 | 20 | -81.0 | |
| *Lincosamide[t]+ToAc[t] | 36 | 21 | 15.8 | 11.7 | 23 | 15.4 | 20 | 0.211 | 29 | 21 | 20 | 21 | -40.0 | 42.1 | 22 | -83.5 | |
| PhCh+PhTh | 24 | 22 | 15.5 | 19.9 | 21 | 15.3 | 21 | 0.211 | 26 | 23 | 21 | 22 | -40.3 | 38.2 | 21 | -83.9 | |
| Retinoid[t] | 5 | 23 | 15.4 | 4.6 | 24 | 15.1 | 25 | 0.205 | 22 | 22 | 25 | 23 | -40.4 | 40.7 | 23 | -84.3 | |
| BP[t]+Lincosamide[t] | 28 | 24 | 15.1 | 5.5 | 22 | 13.0 | 27 | 0.198 | 32 | 24 | 27 | 24 | -40.7 | 40.8 | 24 | -84.9 | |
| *Tetracycline[o]+Comb. PhPe[ph] | 8 | 25 | 13.6 | 14.1 | 26 | 12.9 | 26 | 0.127 | 38 | 25 | 26 | 25 | -42.2 | 42.9 | 25 | -88.1 | |
| Retinoid[t]+Macrolide[t] | 35 | 26 | 13.4 | 10.4 | 28 | 12.8 | 29 | 0.108 | 14 | 27 | 29 | 26 | -42.4 | 41.8 | 26 | -88.2 | |
| Comb. chemical peels[ph] | 14 | 27 | 13.3 | 14.1 | 25 | 12.4 | 32 | 0.089 | 28 | 26 | 32 | 27 | -42.5 | 42.9 | 27 | -88.8 | |
| Benzoyl peroxide[t] | 3 | 28 | 12.8 | 5.4 | 27 | 11.9 | 40 | 0.059 | 37 | 28 | 40 | 28 | -43.0 | 40.7 | 28 | -89.1 | |
| Antiseptics[t] | 8 | 29 | 10.6 | 11.8 | 29 | 9.5 | 38 | 0.045 | 23 | 29 | 38 | 29 | -45.2 | 42.1 | 29 | -93.4 | |

| | | | | | | | | | | | | | | | | |
|---|---|---|---|---|---|---|---|---|---|---|---|---|---|---|---|---|
| Topical acid[t] | 12 | 30 | 9.3 | 8.5 | 30 | 8.8 | 36 | 0.037 | 35 | 32 | 36 | 30 | -46.5 | 41.6 | 30 | -95.8 |
| Macrolide[t] | 7 | 31 | 8.9 | 5.7 | 31 | 8.7 | 30 | 0.035 | 31 | 30 | 30 | 31 | -46.9 | 40.8 | 31 | -96.4 |
| Retinoid tcd<120mg/kg(sc)[o] | 16 | 32 | 8.7 | 12.2 | 32 | 7.0 | 24 | 0.034 | 39 | 31 | 24 | 32 | -47.0 | 41.8 | 32 | -96.9 |
| Co-cyprindiol[o] | 19 | 33 | 7.6 | 8.1 | 34 | 7.0 | 23 | 0.018 | 30 | 33 | 23 | 33 | -48.2 | 41.3 | 33 | -99.0 |
| Comb. Oral Contraceptive[o] | 20 | 34 | 7.3 | 5.7 | 33 | 6.8 | 33 | 0.016 | 36 | 36 | 33 | 34 | -48.5 | 41.0 | 34 | -99.5 |
| Azelaic acid[t] | 6 | 35 | 6.8 | 6.3 | 35 | 6.4 | 28 | 0.012 | 33 | 34 | 28 | 35 | -49.0 | 40.8 | 35 | -100.3 |
| Tetracycline[o] | 17 | 36 | 6.6 | 10.4 | 36 | 4.9 | 31 | 0.003 | 34 | 35 | 31 | 36 | -49.2 | 41.8 | 36 | -101.0 |
| Lincosamide[t] | 4 | 37 | 3.4 | 4.6 | 37 | 2.8 | 35 | 0.002 | 24 | 38 | 35 | 37 | -52.3 | 40.7 | 37 | -106.7 |
| Macrolide[o] | 18 | 38 | 0.7 | 14.4 | 38 | -4.7 | 34 | 0.001 | 20 | 37 | 34 | 38 | -55.1 | 43.0 | 38 | -112.3 |
| Fusidic acid[t] | 9 | 39 | -2.5 | 8.8 | 39 | -7.4 | 39 | 0.001 | 15 | 40 | 39 | 39 | -58.3 | 41.5 | 39 | -118.2 |
| *Anti-fungal[t] | 11 | 40 | -10.0 | 22.3 | 40 | -24.7 | 37 | 0.000 | 17 | 39 | 37 | 40 | -65.8 | 46.0 | 40 | -133.3 |

Abbreviations: BP Benzoyl peroxide; tcd total cumulative dose; [t] topical; [o] oral; [ph] physical; (sc) single course; ToAc Topical Acid; PhCh Photochemical therapy; PhTh Photothermal therapy; PhDy Photodynamic therapy; Comb. Combined; Rk rank (values are ranks on EV)

***Mild-to-Moderate Acne***

***Type of model****:* Random study effects, fixed class effect model with 72 treatments in 40 classes relative to placebo, and bias adjustment for small study effects.

***Outcome***: % Change from Baseline.

***MCID***: 25%, ***GRADE probability cutoff****:* 0.975

***Decision rules Stage 1*** **:**
*EV:* 38 treatment classes have positive EV
*LaEV:* 37 treatment classes have positive LaEV
*GRADE:* only 1 treatment is promoted to Category 1, ranked 2nd on EV.

***Ranking Systems***: Pr(V>T) and Pr(Best) are erratic, respectively privileging less uncertain and more uncertain treatments. However, both Pr(Best) and SUCRA pick out the same top 3 treatments as EV; Pr(V>T) 2 of the 3.

***Decision rules Stage 2***:
*EV*: Only 2 further treatments besides the best treatment are recommended, because they are not worse than the best treatment by more than the MCID.
*LaEV:* Only one further treatment is recommended
*GRADE:* Only the single treatment in Category 1 is recommended

***Comments:*** A number of treatments (marked with an asterisk in Table S2) were excluded from consideration by the guideline development committee on the grounds of small sample size. Two of them would have been recommended by EV. One was excluded by LaEV at Stage 2.

**Fig S2** Mild to Moderate Acne

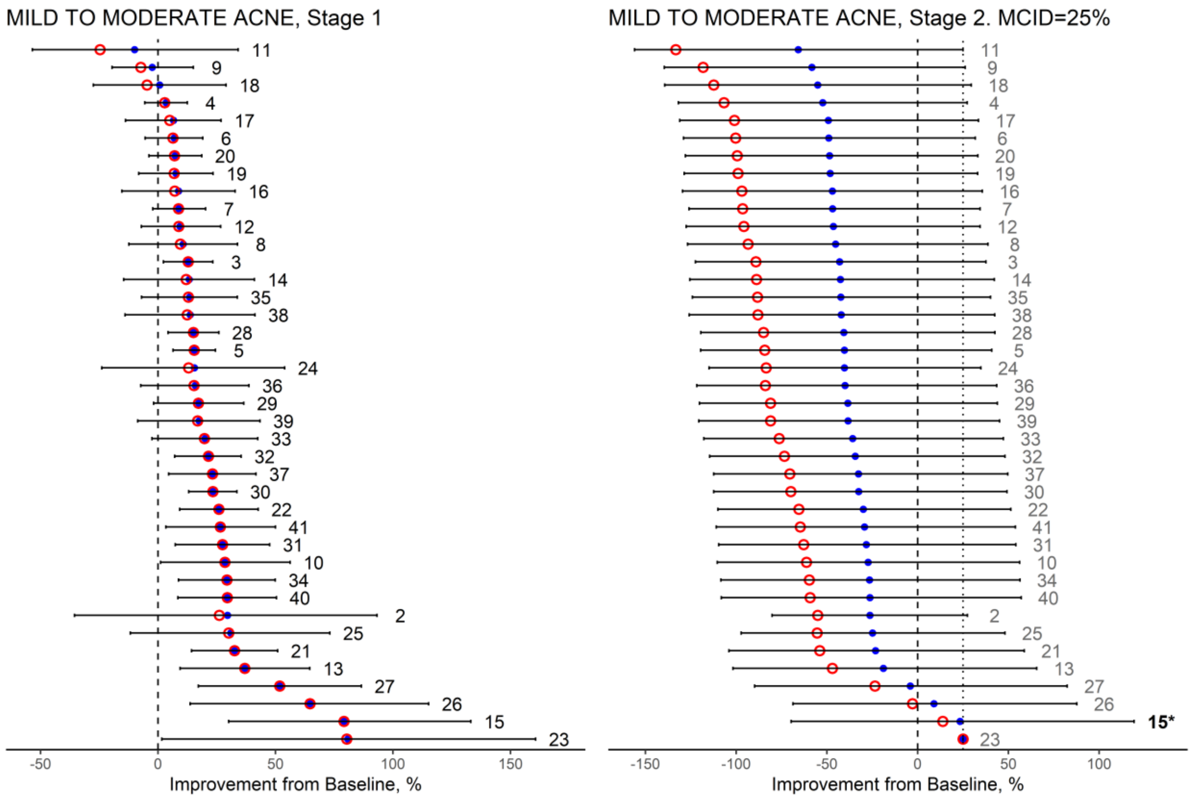

#### A3. Moderate to Severe Depression

**Table S3.**

| Treatment (numbering as in NICE guidelines) | STAGE 1 Decision Rules | | | | | | | Ranking systems | | | STAGE 2 Decision Rules | | | | | |
|---|---|---|---|---|---|---|---|---|---|---|---|---|---|---|---|---|
| | EV | | | LaEV | | GRADE (0.85) Category 1 | | | | | EV | | | LaEV | | GRADE (0.85) Category 1 |
| | Rk | EV | Sd | Rk | LaEV | Rk | Pr(V>T) | P(Best) | SUCRA | Pr(V>T) | Rk | EV | sd | Rk | LaEV | |
| Exercise group + AD 17 | 1 | 1.37 | 0.70 | 1 | 1.36 | 3 | 0.964 | 1 | 1 | 3 | 1(R) | 0.50 | 0 (R) | 1(R) | 0.50 | 3 |
| CT & CBT group + AD 25 | 2 | 1.23 | 0.83 | 2 | 1.20 | 9 | 0.959 | 2 | 3 | 9 | 2 | 0.36 | 1.08 | 3 | 0.14 | 9 |
| CT & CBT individual + AD 6 | 3 | 1.18 | 0.41 | 3 | 1.18 | 1 | 0.903 | 4 | 2 | 1 | 3 | 0.32 | 0.81 | 2 | 0.10 | 1 |
| Yoga group 10 | 4 | 1.05 | 0.61 | 4 | 1.04 | 10 | 0.883 | 3 | 4 | 10 | 4 | 0.19 | 0.92 | 4 | -0.09 | 10 |
| Self-help 21 | 5 | 0.98 | 0.69 | 5 | 0.95 | 6 | 0.857 | 5 | 5 | 6 | 5 | 0.11 | 0.98 | 5 | -0.20 | 6 |
| BT individual 8 | 6 | 0.86 | 0.38 | 6 | 0.86 | 7 | 0.856 | 19 | 6 | 7 | 6 | 0.00 | 0.79 | 6 | -0.31 | 7 |
| Light therapy + AD 11 | 7 | 0.86 | 0.37 | 7 | 0.86 | 5 | 0.847 | 26 | 7 | 5 | 7 | -0.01 | 0.78 | 7 | -0.32 | |
| Problem solving individual 15 | 8 | 0.86 | 0.44 | 8 | 0.85 | 2 | 0.842 | 12 | 8 | 2 | 8 | -0.01 | 0.82 | 8 | -0.34 | |
| Acupuncture + AD 39 | 9 | 0.78 | 0.18 | 9 | 0.78 | 8 | 0.832 | 8 | 9 | 8 | 9 | -0.08 | 0.72 | 9 | -0.40 | |
| CT & CBT individual 13 | 10 | 0.78 | 0.27 | 10 | 0.78 | 4 | 0.832 | 6 | 10 | 4 | 10 | -0.09 | 0.75 | 10 | -0.43 | |
| Counselling individual 14 | 11 | 0.67 | 0.41 | 11 | 0.66 | 11 | 0.699 | 7 | 11 | 11 | 11 | -0.20 | 0.81 | 11 | -0.62 | |
| IPT individual+ AD 22 | 12 | 0.66 | 0.65 | 12 | 0.61 | 13 | 0.634 | 13 | 12 | 13 | 12 | -0.21 | 0.95 | 12 | -0.69 | |
| Self-help with support 19 | 13 | 0.60 | 0.58 | 14 | 0.57 | 12 | 0.598 | 24 | 13 | 12 | 13 | -0.26 | 0.91 | 13 | -0.75 | |
| ST PDT individual 18 | 14 | 0.58 | 0.36 | 13 | 0.56 | 14 | 0.584 | 11 | 14 | 14 | 14 | -0.29 | 0.78 | 14 | -0.76 | |
| IPT individual 16 | 15 | 0.45 | 0.46 | 15 | 0.41 | 15 | 0.450 | 23 | 15 | 15 | 15 | -0.42 | 0.83 | 15 | -1.00 | |
| Acupuncture 24 | 16 | 0.40 | 0.31 | 16 | 0.39 | 19 | 0.415 | 15 | 19 | 19 | 16 | -0.46 | 0.76 | 16 | -1.05 | |
| Mirtazapine 23 | 17 | 0.35 | 0.07 | 17 | 0.35 | 16 | 0.338 | 10 | 16 | 16 | 17 | -0.51 | 0.70 | 17 | -1.12 | |
| SNRIs 4 | 18 | 0.32 | 0.05 | 18 | 0.32 | 26 | 0.266 | 14 | 17 | 26 | 18 | -0.54 | 0.70 | 18 | -1.17 | |
| TCAs 3 | 19 | 0.30 | 0.99 | 20 | 0.29 | 21 | 0.232 | 21 | 18 | 21 | 19 | -0.56 | 1.21 | 20 | -1.24 | |
| ST PDP individual+AD 2 | 20 | 0.29 | 0.12 | 22 | 0.24 | 23 | 0.195 | 9 | 21 | 23 | 20 | -0.58 | 0.70 | 22 | -1.32 | |
| CT & CBT group 12 | 21 | 0.25 | 0.42 | 21 | 0.19 | 24 | 0.147 | 16 | 20 | 24 | 21 | -0.61 | 0.81 | 21 | -1.33 | |
| SSRIs 26 | 22 | 0.24 | 0.04 | 25 | 0.12 | 20 | 0.024 | 20 | 23 | 20 | 22 | -0.63 | 0.69 | 19 | -1.37 | |
| Exercise group 20 | 23 | 0.20 | 0.56 | 23 | 0.10 | 17 | 0.015 | 17 | 26 | 17 | 23 | -0.67 | 0.89 | 23 | -1.45 | |
| Exercise individual 1 | 24 | 0.14 | 0.61 | 19 | 0.05 | 18 | 0.003 | 25 | 24 | 18 | 24 | -0.73 | 0.92 | 25 | -1.53 | |
| Trazodone 9 | 25 | 0.13 | 0.08 | 24 | 0.01 | 22 | 0.000 | 22 | 22 | 22 | 25 | -0.74 | 0.70 | 24 | -1.57 | |
| Individual Counselling+AD 7 | 26 | -0.29 | 1.32 | 26 | -0.96 | 25 | 0.000 | 18 | 25 | 25 | 26 | -1.15 | 1.49 | 26 | -2.49 | |

Abbreviations: R=reference treatment. Rk EV Treatment ranking; AD anti-depressants; CT Cognitive therapy; CBT Cognitive Behavioural therapy; ST Short term; SSRI Selective serotonin reuptake inhibitors ; SNRI Serotonin and norepinephrine reuptake inhibitors ; TCA Tricyclic antidepressant; IPT Interpersonal Psychotherapy; BT Behavioural therapy; PDT Psychodynamic psychotherapy

***Moderate to severe depression***[2]

***Type of model****:* Random study and random class effects model with 99 treatments in 50 treatment classes, relative to placebo, bias-adjusted for small study effects. The committee excluded treatments from the decision set  to which less than 50 patients had been randomized, and other treatments not available in the UK. The decision set consisted of 26 classes of treatment relative to Pill Placebo

***Outcome*****:** Standardized Mean Difference

***MCID***: 0.50  ***GRADE probability cutoff****:* 0.85

***Decision rules Stage 1*** **:**
*EV:* 26 treatment classes had a positive EV
*LaEV:*  The same 26 treatment classes have positive LaEV
*GRADE:*  No treatments were better than Pill Placebo by 0.50 with probability 0.975 or above. The probability threshold 0.85 was selected, and on this basis only 6 treatments were promoted to Category 1, included those ranked 9th and 10th on EV.

***Ranking Systems***: Pr(V>T) ranks treatments 3,9,10 especially highly as these have exceptionally low SD. SUCRA ranking was close to EV; Pr(Best) was highly erratic. However both Pr(Best) and SUCRA pick out the same 5 top-ranked treatments as EV.

***Decision rules Stage 2*****:**
*EV*: Besides the best treatment, a further four treatments are recommended, because they are not worse than the best treatment by more than the MCID.
*LaEV:*  The same four additional treatments are recommended.
*GRADE:* None of the Category 1 treatments is superior to another by the MCID with probably 0.85, so none are promoted to Category 2 and all 6 are recommended

**Figure S3**  Moderate to severe depression

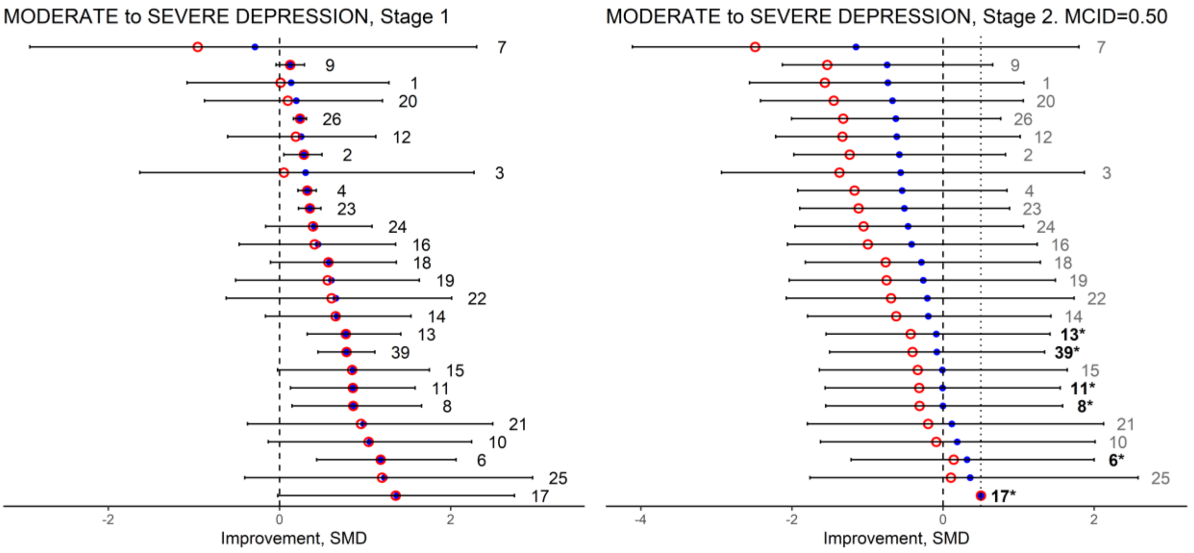

#### A4. Tranexamic Acid (TXA) for primary joint replacement.[3]

**Table S4.** Outcome is reduced risk of transfusion, %, relative to Intra-articular administration

| Treatment (numbering as in NICE guidelines) | STAGE 1 Decision Rules: EV | | | STAGE 1 Decision Rules: LaEV | | STAGE 1 Decision Rules: GRADE (0.975) Category 1 | | Ranking systems | | | STAGE 2 Decision Rules: EV | | | STAGE 2 Decision Rules: LaEV | | STAGE 2 Decision Rules: GRADE (0.975) Category 1 |
|---|---|---|---|---|---|---|---|---|---|---|---|---|---|---|---|---|
| | Rk | EV | Sd | Rk | LaEV | Rk | Pr(V>T) | P(Best) | SUCRA | Pr(V>T) | Rk | EV | sd | Rk | LaEV | |
| Intraarticular and oral (5) | 1 | 6.48 | 4.49 | 1 | 6.33 | 2 | 0.987 | 1 | 1 | 2 | 1(R) | 2.16 | 0 (R) | 1(R) | 2.16 | 2 |
| Intraarticular &Intravenous (4) | 2 | 5.50 | 2.99 | 2 | 5.50 | 1 | 0.933 | 2 | 2 | 1 | 2 | 1.19 | 3.26 | 2 | 0.74 | |
| Oral (3) | 3 | 1.11 | 1.87 | 3 | 0.84 | 3 | 0.169 | 3 | 3 | 3 | 3 | -3.21 | 4.48 | 3 | -6.81 | |
| Intravenous (2) | 4 | 0.55 | 0.99 | 4 | 0.40 | 4 | 0.003 | 4 | 4 | 4 | 4 | -3.77 | 4.40 | 4 | -7.84 | |

##### Tranexamic Acid (TXA) for primary joint replacement.

***Type of model:*** Fixed study, fixed class model with 4 classes of tranexamic acid (TXA) administration relative to Intra-articular administration

***Outcome***: Reduction in risk of transfusion, expressed as %.

***MCID***: RR=1.50. ***GRADE probability cutoff:*** 0.975

***Decision rules Stages 1 and 2*** **:**
*EV and LaEV:* all 4 treatments are superior to the reference treatment EV at Stage 1, and at Stage 2 only one is within an MCID of the best treatment
*GRADE:* Only one treatment, ranked 2$^{nd}$ on EV, was promoted to Category 1, and was therefore recommended.

***Ranking Systems***: All three methods produced the same ranking as EV.

**Figure S4.** Tranexamic Acid in Joint Replacement.

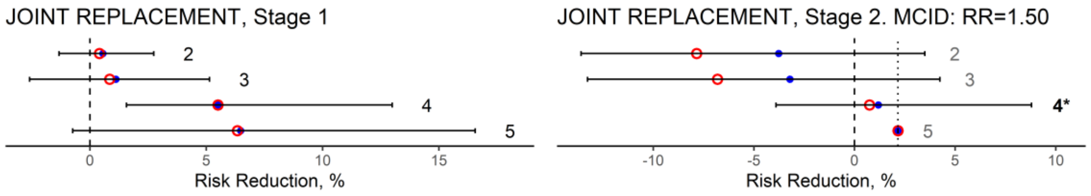

#### A5. Headache Prophylaxis.

**Table S5.** Headache prophylaxis. Outcome is reduction in headache-days per month relative to placebo .

| Treatment (numbering as in NICE guidelines) | STAGE 1 Decision Rules | | | | | | | Ranking systems | | | STAGE 2 Decision Rules | | | | | |
|---|---|---|---|---|---|---|---|---|---|---|---|---|---|---|---|---|
| | EV | | | LaEV | | GRADE (0.975) Category 1 | | | | | EV | | | LaEV | | GRADE (0.975) Category 1 |
| | Rk | EV | Sd | Rk | LaEV | Rk | Pr(V>T) | P(Best) | SUCRA | Pr(V>T) | Rk | EV | sd | Rk | LaEV | |
| Propranolol (6) | 1 | 1.19 | 0.50 | 1 | 1.18 | 3 | 0.988 | 1 | 1 | 3 | 1(R) | 0.50 | 0 (R) | 1(R) | 0.50 | 3 |
| Amitriptyline (2) | 2 | 1.14 | 0.66 | 2 | 1.13 | 1 | 0.925 | 2 | 2 | 1 | 2 | 0.45 | 0.78 | 2 | 0.32 | |
| Topiramate (5) | 3 | 1.04 | 0.24 | 3 | 1.04 | 2 | 0.847 | 3 | 3 | 2 | 3 | 0.35 | 0.48 | 3 | 0.29 | |
| Propranolol & Nadolol (7) | 4 | 0.60 | 0.52 | 4 | 0.57 | 4 | 0.591 | 4 | 4 | 4 | 4 | -0.09 | 0.72 | 4 | -0.41 | |
| Telmisartan (1) | 5 | 0.51 | 0.92 | 5 | 0.35 | 5 | 0.504 | 5 | 5 | 5 | 5 | -0.18 | 1.04 | 5 | -0.68 | |
| Gabapentin (4) | 6 | 0.00 | 0.81 | 6 | -0.32 | 6 | 0.264 | 6 | 6 | 6 | 6 | -0.68 | 0.95 | 6 | -1.50 | |
| Divalproex Sodium (3) | 7 | -0.12 | 0.56 | 7 | -0.40 | 7 | 0.129 | 7 | 7 | 7 | 7 | -0.81 | 0.75 | 7 | -1.67 | |

**Headache prophylaxis**.

***Type of model:*** Random effects model of 7 treatments relative to placebo

***Outcome*****:** Reduction in hospital-days per month

***MCID***: 0.5 days. ***GRADE probability cutoff****:* 0.975

***Decision rules Stages 1*** **& 2**
*EV and LaEV:* At Stage 1, 6 treatments are superior to the placebo on EV and 5 on LaEV. At Stage 2, two treatments were recommended alongside the best treatment; and decisions based on EV and LaEV were identical.
*GRADE:* Only one treatment, the one ranked 3rd on EV, was promoted to Category 1, and was therefore recommended.

***Ranking Systems:*** The three highest-ranked treatments on EV were also ranked highest by SUCRA, Pr(Best) and Pr(V>T).

**Figure S5. Headache prophylaxis**

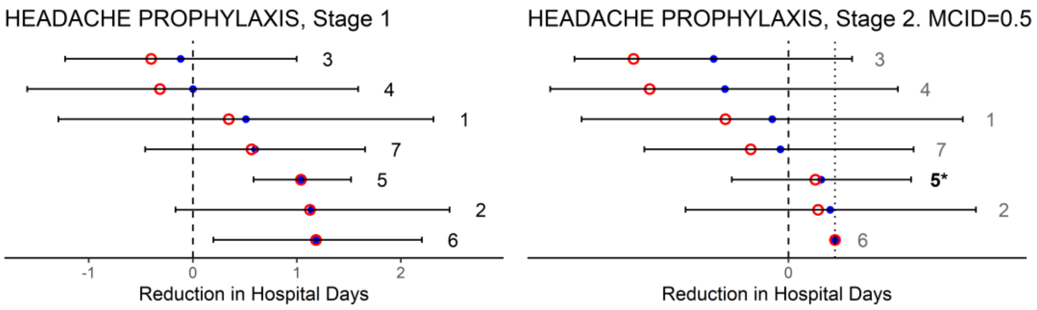

#### A6: Social Anxiety (Treatment) [4]

**Table S6.** Nice Guideline Social Anxiety. Outcome is improvement in clinical score, on SMD scale, relative to waitlist.

| Treatment (numbering as in NICE guidelines) | STAGE 1 Decision Rules | | | | | | | Ranking systems | | | STAGE 2 Decision Rules | | | | | |
|---|---|---|---|---|---|---|---|---|---|---|---|---|---|---|---|---|
| | EV | | | LaEV | | GRADE (0.975) Category 1 | | | | | EV | | | LaEV | | GRADE (0.975) Category 1 |
| | Rk | EV | Sd | Rk | LaEV | Rk | Pr(V>T) | P(Best) | SUCRA | Pr(V>T) | Rk | EV | sd | Rk | LaEV | |
| CBT group Heim + Phenelzine 40 | 1 | 1.68 | 0.21 | 1 | 1.68 | 19 | 1.000 | 1 | 1 | 19 | 1(R) | 0.50 | 0 (R) | 1(R) | 0.50 | 19 |
| Cognitive therapy 35 | 2 | 1.56 | 0.15 | 2 | 1.56 | 5 | 1.000 | 2 | 2 | 5 | 2 | 0.38 | 0.24 | 2 | 0.37 | 5 |
| Paroxetine + Clonazapam 38 | 3 | 1.35 | 0.29 | 3 | 1.35 | 7 | 1.000 | 3 | 5 | 7 | 3 | 0.17 | 0.31 | 3 | 0.11 | 7 |
| Psychodynamic + Clonazepam 37 | 4 | 1.28 | 0.27 | 4 | 1.28 | 2 | 1.000 | 4 | 3 | 2 | 4 | 0.10 | 0.30 | 5 | 0.05 | 2 |
| Phenelzine 22 | 5 | 1.27 | 0.15 | 5 | 1.27 | 1 | 1.000 | 6 | 4 | 1 | 5 | 0.09 | 0.20 | 4 | 0.02 | 1 |
| CBT individual + Moclobemide 39 | 6 | 1.23 | 0.25 | 6 | 1.23 | 11 | 1.000 | 28 | 7 | 11 | 6 | 0.05 | 0.29 | 6 | -0.04 | 11 |
| CBT individual 34 | 7 | 1.19 | 0.15 | 7 | 1.19 | 22 | 1.000 | 8 | 6 | 22 | 7 | 0.00 | 0.24 | 7 | -0.09 | 22 |
| CBT group enhanced exposure 31 | 8 | 1.10 | 0.20 | 8 | 1.10 | 12 | 1.000 | 30 | 8 | 12 | 8 | -0.08 | 0.27 | 8 | -0.24 | 12 |
| Clonazapam 20 | 9 | 1.07 | 0.19 | 9 | 1.07 | 13 | 0.999 | 5 | 9 | 13 | 9 | -0.11 | 0.25 | 9 | -0.28 | 13 |
| CBT individual Heimberg 33 | 10 | 1.02 | 0.20 | 10 | 1.02 | 8 | 0.999 | 10 | 11 | 8 | 10 | -0.16 | 0.29 | 10 | -0.37 | 8 |
| Paroxetine 17 | 11 | 0.99 | 0.14 | 11 | 0.99 | 9 | 0.999 | 18 | 10 | 9 | 11 | -0.19 | 0.21 | 11 | -0.40 | 9 |
| CT abbreviated experiments 32 | 12 | 0.97 | 0.12 | 12 | 0.97 | 3 | 0.998 | 7 | 12 | 3 | 12 | -0.21 | 0.23 | 12 | -0.45 | 3 |
| Venlafaxine <75 18 | 13 | 0.96 | 0.15 | 13 | 0.96 | 6 | 0.998 | 17 | 13 | 6 | 13 | -0.22 | 0.21 | 13 | -0.46 | 6 |
| CBT group + Fluoxetine 36 | 14 | 0.95 | 0.19 | 14 | 0.95 | 4 | 0.997 | 23 | 14 | 4 | 14 | -0.23 | 0.26 | 14 | -0.49 | 4 |
| Fluvoxamine 16 | 15 | 0.94 | 0.16 | 15 | 0.94 | 25 | 0.997 | 9 | 15 | 25 | 15 | -0.25 | 0.22 | 15 | -0.51 | 25 |
| Sertraline 12 | 16 | 0.91 | 0.16 | 16 | 0.91 | 15 | 0.997 | 39 | 16 | 15 | 16 | -0.27 | 0.22 | 16 | -0.55 | 15 |
| Gabapentin 10 | 17 | 0.89 | 0.27 | 17 | 0.89 | 27 | 0.996 | 14 | 17 | 27 | 17 | -0.30 | 0.31 | 17 | -0.62 | 27 |
| Social skills training 24 | 18 | 0.88 | 0.25 | 18 | 0.88 | 29 | 0.996 | 26 | 19 | 29 | 18 | -0.31 | 0.32 | 19 | -0.62 | 29 |
| Self-help internet with support 7 | 19 | 0.88 | 0.08 | 19 | 0.88 | 16 | 0.996 | 24 | 18 | 16 | 19 | -0.31 | 0.22 | 20 | -0.62 | 16 |
| Escitalopram 14 | 20 | 0.88 | 0.16 | 20 | 0.88 | 10 | 0.994 | 12 | 20 | 10 | 20 | -0.31 | 0.23 | 18 | -0.64 | 10 |
| Fluoxetine 15 | 21 | 0.87 | 0.15 | 21 | 0.87 | 21 | 0.992 | 36 | 21 | 21 | 21 | -0.32 | 0.23 | 21 | -0.64 | 21 |
| CBT group 29 | 22 | 0.85 | 0.09 | 22 | 0.85 | 14 | 0.990 | 34 | 23 | 14 | 22 | -0.33 | 0.21 | 22 | -0.67 | 14 |
| Alprazolam 19 | 23 | 0.85 | 0.28 | 24 | 0.85 | 20 | 0.990 | 25 | 24 | 20 | 23 | -0.33 | 0.26 | 24 | -0.68 | 20 |
| Self-help book with support 6 | 24 | 0.85 | 0.16 | 23 | 0.85 | 24 | 0.983 | 33 | 22 | 24 | 24 | -0.33 | 0.32 | 23 | -0.69 | 24 |
| Self-help book no support 4 | 25 | 0.84 | 0.12 | 25 | 0.84 | 31 | 0.943 | 19 | 25 | 31 | 25 | -0.34 | 0.24 | 25 | -0.69 | |
| Citalopram 13 | 26 | 0.83 | 0.23 | 26 | 0.83 | 18 | 0.933 | 32 | 28 | 18 | 26 | -0.35 | 0.28 | 26 | -0.71 | |
| Exposure in vivo 23 | 27 | 0.83 | 0.12 | 27 | 0.83 | 26 | 0.930 | 16 | 26 | 26 | 27 | -0.36 | 0.23 | 27 | -0.72 | |
| Levetiracetam 9 | 28 | 0.83 | 0.33 | 28 | 0.83 | 17 | 0.929 | 20 | 27 | 17 | 28 | -0.36 | 0.37 | 28 | -0.75 | |
| CBT group Heimberg 30 | 29 | 0.80 | 0.11 | 29 | 0.80 | 23 | 0.895 | 21 | 30 | 23 | 29 | -0.38 | 0.20 | 29 | -0.77 | |
| Mirtazapine 11 | 30 | 0.80 | 0.33 | 30 | 0.80 | 32 | 0.891 | 15 | 29 | 32 | 30 | -0.38 | 0.36 | 30 | -0.79 | |

| | | | | | | | | | | | | | | | |
|---|---|---|---|---|---|---|---|---|---|---|---|---|---|---|---|
| Moclobemide 21 | 31 | 0.73 | 0.15 | 31 | 0.73 | 33 | 0.879 | 11 | 31 | 33 | 31 | -0.45 | 0.22 | 31 | -0.90 |
| Pregabalin 400 8 | 32 | 0.72 | 0.18 | 32 | 0.72 | 28 | 0.839 | 13 | 32 | 28 | 32 | -0.46 | 0.24 | 32 | -0.93 |
| Self-help internet no support 5 | 33 | 0.66 | 0.14 | 33 | 0.66 | 34 | 0.824 | 31 | 33 | 34 | 33 | -0.52 | 0.25 | 33 | -1.04 |
| Attention-matched control 2 | 34 | 0.63 | 0.14 | 34 | 0.63 | 30 | 0.820 | 27 | 35 | 30 | 34 | -0.55 | 0.23 | 34 | -1.11 |
| Psychodynamic psychotherapy 28 | 35 | 0.62 | 0.16 | 35 | 0.62 | 35 | 0.778 | 40 | 34 | 35 | 35 | -0.56 | 0.25 | 35 | -1.13 |
| Pill Placebo 1 | 36 | 0.47 | 0.12 | 36 | 0.47 | 36 | 0.392 | 38 | 39 | 36 | 36 | -0.72 | 0.20 | 36 | -1.43 |
| Interpersonal psychotherapy 27 | 37 | 0.43 | 0.20 | 37 | 0.43 | 37 | 0.366 | 37 | 37 | 37 | 37 | -0.75 | 0.28 | 37 | -1.50 |
| Mindfulness cognitive therapy 26 | 38 | 0.39 | 0.22 | 38 | 0.39 | 39 | 0.338 | 35 | 36 | 39 | 38 | -0.79 | 0.28 | 38 | -1.58 |
| Exercise 3 | 39 | 0.35 | 0.37 | 39 | 0.31 | 38 | 0.309 | 22 | 38 | 38 | 39 | -0.83 | 0.41 | 39 | -1.67 |
| Supportive therapy 25 | 40 | 0.26 | 0.24 | 40 | 0.24 | 40 | 0.152 | 29 | 40 | 40 | 40 | -0.92 | 0.30 | 40 | -1.85 |

**Social Anxiety (Treatment)**

***Type of model****:* Random study effects, random class model (17 classes), with 40 individual treatments relative to waitlist control forming the decision set.[4,5]

***Outcome*****:** Standardized Mean Difference

***MCID***: 0.50  ***GRADE probability cutoff****:* 0.975

***Decision rules Stage 1*** **:**
*EV and LaEV.* All 40 treatments were superior to waitlist control.
*GRADE:* 24 treatments were promoted to Category 1.

***Ranking Systems****:* SUCRA picks out the same top 7 treatments as EV. Pr(V>T) accords the highest rank to the treatment ranked 19th by EV and LaEV, due to its exceptionally low SD. Pr(Best) places the treatments ranked 28th by EV and LaEV in the top 7 treatments, due to their exceptionally high SD.

***Decision rules Stage 2***
*EV*: Besides the best treatment, a further 6 treatments are recommended, because they are not worse than the best treatment by more than the MCID.
*LaEV:* Only four of the six additional treatments are recommended by LaEV
*GRADE:* None of the Category 1 treatments is superior to another by the MCID with probability 0.975, so none are promoted to Category 2 and all 24 are recommended

**Figure S6.** Social Anxiety (Treatment).

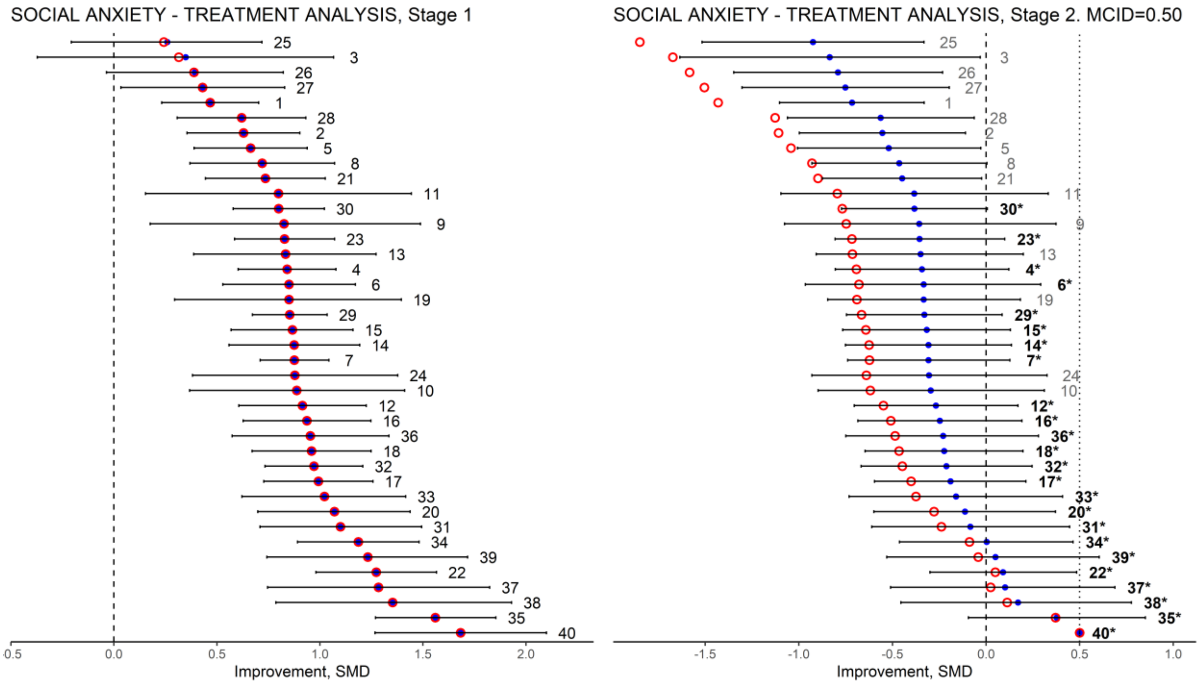

#### A7. Social Anxiety, Class analysis[6]

**Table S7.** Social Anxiety, class analysis. . Outcome is improvement in clinical score, on SMD scale, relative to waitlist.

| Treatment (numbering as in NICE guidelines) | STAGE 1 Decision Rules | | | | | | | Ranking systems | | | STAGE 2 Decision Rules | | | | | |
|---|---|---|---|---|---|---|---|---|---|---|---|---|---|---|---|---|
| | EV | | | LaEV | | GRADE (0.975) Category 1 | | | | | EV | | | LaEV | | GRADE (0.975) Category 1 |
| | Rk | EV | Sd | Rk | LaEV | Rk | Pr(V>T) | P(Best) | SUCRA | Pr(V>T) | Rk | EV | sd | Rk | LaEV | |
| Combined 17 | 1 | 1.30 | 0.22 | 1 | 1.30 | 1 | 1.000 | 1 | 1 | 1 | 1(R) | 0.50 | 0 (R) | 1(R) | 0.50 | 1 |
| CBT individual 16 | 2 | 1.19 | 0.19 | 2 | 1.19 | 2 | 0.999 | 2 | 2 | 2 | 2 | 0.38 | 0.27 | 2 | 0.37 | 2 |
| MAOI 11 | 3 | 1.00 | 0.28 | 3 | 1.00 | 6 | 0.995 | 3 | 3 | 6 | 3 | 0.20 | 0.33 | 3 | 0.15 | 6 |
| Benzodiazepines 10 | 4 | 0.96 | 0.31 | 4 | 0.96 | 5 | 0.977 | 10 | 4 | 5 | 4 | 0.16 | 0.34 | 4 | 0.09 | 5 |
| CBT group 15 | 5 | 0.92 | 0.21 | 5 | 0.92 | 3 | 0.963 | 4 | 5 | 3 | 5 | 0.12 | 0.28 | 6 | 0.06 | |
| SSRI/SNRI 9 | 6 | 0.91 | 0.16 | 6 | 0.91 | 4 | 0.935 | 8 | 6 | 4 | 6 | 0.11 | 0.23 | 5 | 0.05 | |
| Self-help with support 6 | 7 | 0.86 | 0.25 | 7 | 0.86 | 7 | 0.930 | 7 | 7 | 7 | 7 | 0.06 | 0.33 | 7 | -0.04 | |
| Exposure 12 | 8 | 0.85 | 0.28 | 8 | 0.85 | 8 | 0.899 | 5 | 8 | 8 | 8 | 0.05 | 0.35 | 8 | -0.06 | |
| Anticonvulsants 7 | 9 | 0.81 | 0.27 | 9 | 0.81 | 9 | 0.877 | 13 | 9 | 9 | 9 | 0.01 | 0.32 | 9 | -0.11 | |
| NSSA 8 | 10 | 0.80 | 0.42 | 10 | 0.79 | 11 | 0.851 | 9 | 10 | 11 | 10 | 0.00 | 0.45 | 10 | -0.18 | |
| Self-help no support 5 | 11 | 0.75 | 0.25 | 11 | 0.75 | 12 | 0.824 | 16 | 11 | 12 | 11 | -0.05 | 0.33 | 11 | -0.21 | |
| Psychological placebo 3 | 12 | 0.63 | 0.14 | 12 | 0.63 | 10 | 0.765 | 11 | 13 | 10 | 12 | -0.17 | 0.23 | 12 | -0.38 | |
| Psychodynamic psychotherapy 14 | 13 | 0.62 | 0.35 | 13 | 0.61 | 13 | 0.641 | 14 | 12 | 13 | 13 | -0.18 | 0.41 | 13 | -0.45 | |
| Pill placebo 2 | 14 | 0.47 | 0.37 | 14 | 0.45 | 14 | 0.461 | 6 | 14 | 14 | 14 | -0.33 | 0.40 | 14 | -0.71 | |
| Other psychological therapies 13 | 15 | 0.36 | 0.25 | 15 | 0.35 | 16 | 0.376 | 15 | 16 | 16 | 15 | -0.44 | 0.31 | 15 | -0.89 | |
| Exercise promotion 4 | 16 | 0.35 | 0.49 | 16 | 0.28 | 15 | 0.280 | 12 | 15 | 15 | 16 | -0.45 | 0.53 | 16 | -0.96 | |

***Social Anxiety, Class analysis***

***Type of model****:* Random study effects, random class model: 16 treatment classes based on 40 individual treatments, with waitlist as the reference.[4,5]

***Outcome*****:** Standardized Mean Difference

***MCID***: 0.50 ***GRADE probability cutoff****:* 0.975

***Decision rules Stage 1*** **:**
*EV and LaEV.* All 16 treatment classes were superior to waitlist control.
*GRADE:* 4 treatments were promoted to Category 1.

***Ranking Systems****:* Both Pr(Best) and SUCRA pick out the same 9 top-ranked treatments as EV, and Pr(V>T) identifies 2 of them.

***Decision rules Stage 2*****:**
*EV*: Besides the best treatment, a further 8 treatment classes are recommended, because they are not worse than the best treatment by more than the MCID.
*LaEV:* Only 5 of the 8 additional treatments are recommended by LaEV.
*GRADE:* None of the four Category 1 treatments is superior to another by the MCID with probably 0.975, so none are promoted to Category 2 . All 4 are recommended.

**Figure S7.** Social Anxiety (Class Analysis)

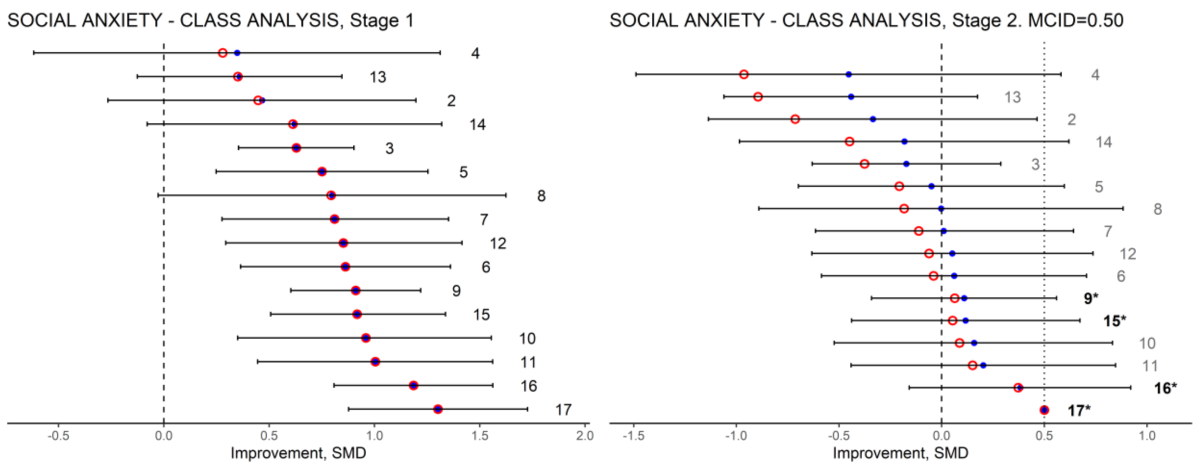

#### A8. Urinary Incontinence

T**able S8.** Urinary incontinence. The outcome is probability of achieving continence status, relative to placebo.

| Treatment (numbering as in NICE guidelines) | STAGE 1 Decision Rules | | | | | | | Ranking systems | | | STAGE 2 Decision Rules | | | | | |
|---|---|---|---|---|---|---|---|---|---|---|---|---|---|---|---|---|
| | EV | | | LaEV | | GRADE (0.975) Category 1 | | | | | EV | | | LaEV | | GRADE (0.975) Category 1 |
| | Rk | EV | Sd | Rk | LaEV | Rk | Pr(V>T) | P(Best) | SUCRA | Pr(V>T) | Rk | EV | sd | Rk | LaEV | |
| Oxybutynin IR (1) | 1 | 0.124 | 0.038 | 1 | 0.124 | 7 | 1.000 | 1 | 1 | 7 | 1(R) | 0.043 | 0 (R) | 1(R) | 0.043 | 7 |
| Tolterodine IR (4) | 2 | 0.116 | 0.053 | 2 | 0.116 | 11 | 1.000 | 2 | 2 | 11 | 2 | 0.035 | 0.051 | 2 | 0.028 | 11 |
| Darifenacin (11) | 3 | 0.098 | 0.052 | 3 | 0.098 | 8 | 1.000 | 4 | 3 | 8 | 3 | 0.017 | 0.071 | 3 | -0.003 | 8 |
| Propiverine IR (5) | 4 | 0.094 | 0.070 | 4 | 0.094 | 13 | 0.996 | 3 | 4 | 13 | 4 | 0.012 | 0.084 | 4 | -0.014 | 13 |
| Trospium (9) | 5 | 0.084 | 0.047 | 5 | 0.084 | 6 | 0.993 | 9 | 5 | 6 | 5 | 0.002 | 0.067 | 5 | -0.023 | 6 |
| Oxybutynin ER (3) | 6 | 0.079 | 0.038 | 6 | 0.079 | 3 | 0.989 | 5 | 6 | 3 | 6 | -0.002 | 0.060 | 6 | -0.027 | 3 |
| Solifenacin (2) | 7 | 0.071 | 0.025 | 7 | 0.071 | 1 | 0.985 | 6 | 7 | 1 | 7 | -0.011 | 0.053 | 7 | -0.038 | 1 |
| Trospium ER (12) | 8 | 0.070 | 0.028 | 8 | 0.070 | 10 | 0.982 | 10 | 8 | 10 | 8 | -0.011 | 0.054 | 8 | -0.039 | 10 |
| Oxybutynin TG (13) | 9 | 0.068 | 0.062 | 9 | 0.067 | 2 | 0.973 | 8 | 9 | 2 | 9 | -0.013 | 0.077 | 9 | -0.050 | |
| Oxybutynin TD (10) | 10 | 0.064 | 0.031 | 10 | 0.064 | 5 | 0.970 | 12 | 10 | 5 | 10 | -0.018 | 0.055 | 10 | -0.051 | |
| Fesoterodine (8) | 11 | 0.057 | 0.020 | 11 | 0.057 | 12 | 0.953 | 7 | 11 | 12 | 11 | -0.025 | 0.049 | 11 | -0.060 | |
| Propiverine ER (7) | 12 | 0.055 | 0.029 | 12 | 0.055 | 4 | 0.901 | 11 | 12 | 4 | 12 | -0.027 | 0.053 | 12 | -0.064 | |
| Tolterodine ER (6) | 13 | 0.036 | 0.014 | 13 | 0.036 | 9 | 0.793 | 13 | 13 | 9 | 13 | -0.045 | 0.045 | 13 | -0.095 | |

#### *Urinary Incontinence*

***Type of model:*** Fixed study effects, 13 treatments compared to placebo.[7,8]

***Outcome*:** Probability of improvement in continence status

***MCID***: RR=1.25 ***GRADE probability cutoff***: 0.975

***Decision rules Stage 1*** **:**
*EV and LaEV.* All 13 treatments were superior to placebo.
*GRADE:* 8 treatments were promoted to Category 1.

***Ranking Systems***: SUCRA top-ranked the same 5 treatments as EV, Pr(best) 4 of them. Pr(V>T) selected none of them, but privileged treatments 11 and 13 which had exceptionally certain effects.

***Decision rules Stage 2***
*EV*: Besides the best treatment, a further 4 treatments were recommended, because they are not worse than the best treatment by more than the MCID.
*LaEV:* Only 1 further treatment recommended besides the best treatment.
*GRADE:* None of the Category 1 treatments is superior to any other by the MCID with probably 0.975, so none are promoted to Category 2, and all 8 are recommended.

**Comments**: The results with LaEV concur with a threshold analysis in which a plausible degree of bias (0.1 on the log odds scale) in the evidence for oxybutynin IR would make tolterodine IR the best treatment.[8] A further point of interest in this NMA is that the guideline developers made oxybutynin IR the reference treatment, rather than placebo, from the outset. This was because it had been shown to be the best treatment in an earlier evaluation. This suggests that the 2-stage methodology, in which treatments are compared to the best treatment in the second stage, might be readily adopted by clinical decision makers.

**Figure S8** Urinary Incontinence

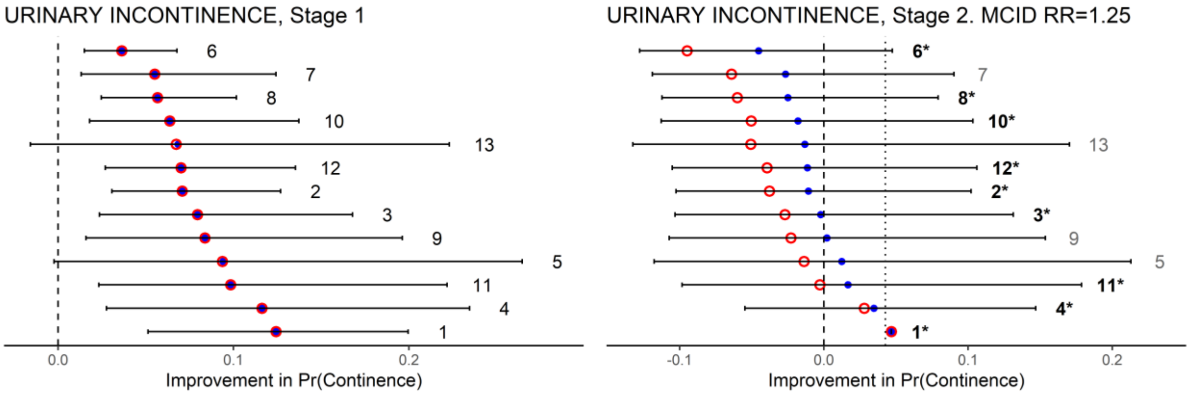

#### A9. Tocolytic therapy in Preterm Labour.[8,9]

**Table S9.** Tocolytic therapy in preterm Labour. Outcome is increase in EGA (weeks)

| **Treatment** (numbering as in NICE guidelines) | **STAGE 1 Decision Rules** | | | | | | | **Ranking systems** | | | **STAGE 2 Decision Rules** | | | | | |
|---|---|---|---|---|---|---|---|---|---|---|---|---|---|---|---|---|
| | **EV** | | | **LaEV** | | **GRADE** (0.975) Category 1 | | | | | **EV** | | | **LaEV** | | **GRADE** (0.975) Category 1 |
| | **Rk** | **EV** | **Sd** | **Rk** | **LaEV** | **Rk** | **Pr(V>T)** | **P(Best)** | **SUCRA** | **Pr(V>T)** | **Rk** | **EV** | **sd** | **Rk** | **LaEV** | |
| Prostaglandin inhibs. (2) | 1 | 2.32 | 0.53 | 1 | 2.32 | 1 | 0.992 | 1 | 1 | 1 | 1(R) | 1.00 | 0 (R) | 1(R) | 1.00 | 1 |
| Ca channel blockers (5) | 2 | 1.68 | 0.50 | 2 | 1.68 | 2 | 0.915 | 3 | 2 | 2 | 2 | 0.37 | 0.53 | 2 | 0.29 | |
| Nitrates (6) | 3 | 1.65 | 0.57 | 3 | 1.65 | 3 | 0.875 | 2 | 3 | 3 | 3 | 0.34 | 0.67 | 3 | 0.21 | |
| Magnesium sulphate (3) | 4 | 1.28 | 0.50 | 4 | 1.28 | 5 | 0.723 | 6 | 4 | 5 | 4 | -0.03 | 0.50 | 4 | -0.25 | |
| Betamimetics (4) | 5 | 1.24 | 0.42 | 5 | 1.24 | 4 | 0.716 | 4 | 5 | 4 | 5 | -0.07 | 0.52 | 5 | -0.31 | |
| Oxytocin RBs (7) | 6 | 0.67 | 1.02 | 6 | 0.52 | 6 | 0.371 | 5 | 6 | 6 | 6 | -0.64 | 1.09 | 6 | -1.47 | |

Abbreviations: inhibs. Inhibitors; Ca Calcium; RBs receptor inhibitors

***Tocolytic therapy in preterm Labour***

***Type of model**:* Random study effects, fixed class NMA of 6 treatment classes compared to placebo.

***Outcome***: Increase in Expected Gestational Age (EGA) at delivery, measured in weeks.

***MCID***: 1 week ***GRADE probability cutoff***: 0.975

***Decision rules Stage 1* & 2:**
*EV and LaEV.* All 6 treatments were superior to placebo, and 2 treatments had EGA within 1 week of the best treatment. Recommendations for EV and LaEV were the same
*GRADE:* A single treatment is promoted to Category 1, and is therefore recommended, the same treatment was top-ranked by EV.

***Ranking systems**:* The three treatments top-ranked on EV and LaEV are also top-ranked by all three methods

**Figure S9**. Tocolytic therapy in preterm Labour

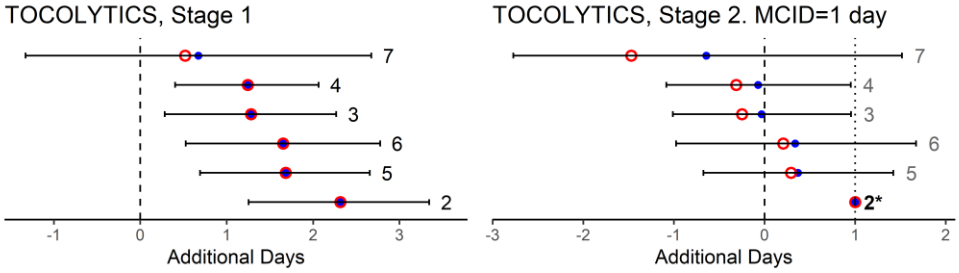

### APPENDIX B: WinBUGS CODE for ILLUSTRATIVE EXAMPLES

#### ILLUSTRATION 1:

```
model {
 for (s in 1:100)  {  p[s] <- pow(s/20,-2)
                              v[s] ~ dnorm(1,p[s])
                              el[s] <- max(0,-v[s])
                              laev[s] <- 1-el[s]
                              ev[s] <- 1.0
                              pr[1,s] <- step(v[s]-0)
                              pr[2,s] <- step(v[s]-0.5)
                              pr[3,s] <- step(v[s]-1.0)
                              pr[4,s] <- step(v[s]-1.5)
                              pr[5,s] <- step(v[s]-2.0)
                              pr[6,s] <- step(v[s]-3.0)  }
}
```

#### ILLUSTRATION 2:

```
model {
 for (s in 1:3)  {  p[s] <- pow(sd[s],-2)
                    v[s] ~ dnorm(m[s],p[s])
                    el[s] <- max(0,-v[s])
                    laev[s] <- v[s]-el[s] }
          }

list(m=c(1,2,3), sd=c(0.1,1,1.5))
```

#### ILLUSTRATION 4:

```
model {

for (i in 1:nm) { for (j in 1:nsd) {
                          m[i,j] <- 1+i/10                  # generates EVs at 1.1, 1.2, 1.3, 1.4, 1.5
                          sd[i,j] <- sdlist[j]
                          pr[i,j] <- pow(sd[i,j],-2)
                          t[i,j] ~ dnorm(m[i,j],pr[i,j])
                          ev[i,j] <- t[i,j]                 # incremental benefit relative to standard
                          el[i,j] <- max(0,-ev[i,j])        # exp incr value if perfect info
                          laev[i,j] <- ev[i,j] - el[i,j]    # loss-adjusted expected value
                          pv[1,i,j] <- step(t[i,j]-0.6)
                          pv[2,i,j] <- step(t[i,j]- 1.3)
                          pv[3,i,j] <- step(t[i,j]-2.0)
                          tx[(i-1)*nsd + j] <- t[i,j] }}

for (i in 1:25) {  rk[i] <- 26 - rank(tx[],i)                                        # mean rank (check)
                   pb[i] <- equals(rk[i],1)                                          # probability best
                   for (j in 1:25) { pbij[i,j] <- step(tx[i]-tx[j]) - equals(i,j)  }  # count if (i > j)
                   su[i] <- sum(pbij[i,]) / 24 }                                     # SUCRA
}

list(nm=5, nsd=5,  sdlist=c(1,2,3,4,5) )
```

### APPENDIX C: WINBUGS CODE FOR NMAs

##### Additional code for Social anxiety Class analysis, 16 treatments

```
#  0.5 SMD as the MCID
# STAGE 1
for (i in 1:16) {v[i] <- -m[i+1]                 # EV
                la[i] <- v[i] - max(0,-v[i])      # LaEV
                pv[i] <- step(v[i] - 0.5)         # Pr(V>T)
                rnk[i] <- 17-rank(v[],i)          # SUCRA
                bst[i] <- equals(rnk[i],1)}       # Pr(Best)

# STAGE 2 (treatment 16 is best)
for (i in 1:15) { v2[i] <- 0.50 - (v[16]-v[i])    # EV
                 la2[i] <- v2[i] - max(0,-v2[i])} # LaEV

# GRADE
for (i in 1:16) {for (j in i+1:17) { v3[j,i] <- -m[j]+ m[i]
                                     gr2[j,i] <- step(v3[j,i]-0.5) }}
for (j in 1:16) {for (i in j+1:17)  { v3[j,i] <- -m[j]+ m[i]
                                     gr2[j,i] <- step(v3[j,i]-0.5) }}
```

#### Supplementary Material References

1. National Institute for Health and Care Excellence. Acne vulgaris: management. NICE Guideline [NG 198]. London, 2021.
2. National Institute for Health and Care Excellence. Depression in adults: treatment and management. NICE Guideline [NG 222]. London, 2022.
3. National Institute for Health and Clinical Excellence. Joint replacement (primary): hip, knee and shoulder. Network meta-analysis and cost analysis of methods for tranexamic acid administration. NG157. London, 2020.
4. National Institute for Health and Clinical Excellence. Social Anxiety Disorder: Recognition, Assessment and Treatment. CG159. London, 2020.
5. Phillippo DM, Dias S, Welton NJ, Caldwell DC, Taske N, Ades AE. Threshold Analysis as an Alternative to GRADE for Assessing Confidence in Guideline Recommendations Based on Network Meta-analyses. *Ann Intern Med* 2019; **170**: 538-46.
6. Mayo-Wilson E, Dias S, Mavranezouli I, et al. Psychological and pharmacological interventions for social anxiety disorder in adults: a systematic review and network meta-analysis. *Lancet Psychiatry* 2014; **1**: 368-76.
7. National Institute for Health and Clinical Excellence. Urinary incontinence in women: the management of urinary incontinence in women. London, 2013.
8. Phillippo DM, Dias S, Welton NJ, Ades AE. Threshold Analysis in NICE Guideline Development, https://www.bristol.ac.uk/population-health-sciences/centres/cresyda/mpes/nice/tsu-reports/, 2016.
9. National Institute for Health and Clinical Excellence. Preterm labour and birth. London, 2015.
